# Pathogenic Epilepsy Gene Variant Prevalence and Penetrance Among U.S. Military Veterans in the Million Veteran Program Cohort

**DOI:** 10.64898/2026.08.18.26360604

**Authors:** Marissa Kellogg, Andrea Hildebrand, Tia Dinatale, Manaz Rezayee, Jessica Minnier, Andrea L.C. Schneider, Lia Ernst, Michelle Cameron, Elizabeth Gerard, Alica Goldman, Remi Stevelink, Amy Brooks-Kayal, Kathryn M. Pridgen, VA Million Veteran Program, Julie A. Lynch, Craig C. Teerlink

## Abstract

**Background and Objectives:** Genetic causes of epilepsy are well-established in children, but the genetics of adult-onset epilepsy is not well understood. There are few studies of epilepsy genetics in older adults, U.S. military Veterans, and people with acquired causes of epilepsy like traumatic brain injury (TBI) and stroke. To test if rare gene variants that cause pediatric epilepsy are associated with adult-onset epilepsy, we determined the prevalence of pathogenic germline variants (PGVs) in epilepsy-associated genes in an ancestrally diverse cohort of older Veterans and examined the penetrance of epilepsy among PGV carriers. We evaluated the effect of mode of inheritance (MOI), variant selection, single gene-level factors, and gene-disease relationship validity on prevalence and penetrance estimates.

**Methods:** This retrospective cohort study used electronic health record (EHR) data from Veterans enrolled in the Million Veteran Program (MVP) biobank who had whole genome sequencing (WGS) data available. We identified Veterans with one or more rare (variant allele frequency [VAF] <0.01) pathogenic/likely pathogenic single nucleotide variants (SNVs) within one or more of 165 expert-curated epilepsy genes. Epilepsy phenotype was defined using a validated algorithm, and penetrance estimates were calculated using Bayes theorem and compared to civilian cohorts.

**Results:** There were 102,624 MVP participants with WGS data. Mean age at censorship or death was 74.6 years, 6.1% were female and 6.3% had epilepsy. Among participants, 1.9% (n=1,955) carried at least 1 rare PGVs and 1.0% (n=1,041) carried ultrarare PGVs. Most carriers of autosomal dominant (AD) PGVs (89.7%) were not diagnosed with epilepsy, though carriers of both AD and autosomal recessive (AR) ultrarare PGVs had increased odds of epilepsy (odds ratios of 1.72 and 1.45, respectively) compared to non-carriers. Penetrance estimates were low for AD PGVs (8.2%), but similar to estimates from civilian biobanks.

**Discussion:** Veterans carrying PGVs in AD-labeled epilepsy genes had increased risk for epilepsy, but only 10.3% were diagnosed. Unexpectedly, Veterans heterozygous for AR-labeled PGVs also had increased risk of epilepsy. Potential reasons for this include latent compound heterozygosity, misclassification of variant pathogenicity or gene MOI, or the possibility that PGVs in AR genes may be risk alleles for adult-onset epilepsy.

## Introduction

Epilepsy is a common neurological condition marked by recurrent seizures with estimated U.S.-based prevalences of 0.7-1.1% in adults^1–4^, 3.0% in elderly adults^5^, and 0.6-2.9% in U.S. military Veterans^2,6,7^. The higher epilepsy prevalence in Veterans is partially explained by the increased prevalence of traumatic brain injury (TBI)^2,7^ and stroke^8,9^. Over 1,000 epilepsy-associated genes^10^ have been discovered; however, inheritance of epilepsy is rarely classically Mendelian, and phenotype-genotype relationships are complex. Most epilepsy genetics research and gene discovery has been conducted in children and young adults of European ancestry, excluding individuals with late-onset or acquired epilepsy due to TBI or stroke^11^, and no studies have been performed in U.S. Veterans. The few studies in adults have typically focused on polygenic risk^12,13^. The effect of rare pathogenic variants (PGVs) in epilepsy genes on adult-onset epilepsy is not well understood^14^.

Recent largescale biobank studies report unexpectedly low penetrance of monogenic disorders among carriers of pathogenic variants in autosomal dominant (AD) genes^15,16^, including genes associated with epilepsy and developmental epileptic encephalopathy (DEE)^3,17^. While family-based studies typically describe penetrance ranging from 20-100% for most epilepsy and DEE-associated genes, Stevelink et al.^3^ found childhood-onset epilepsy penetrance was between 4.1-9.8% in carriers of ultrarare PGVs from the U.K. Biobank (UKB)^18^ and Genomics England (GEL) cohorts. These findings highlight the differences between penetrance estimates derived from family-based studies of disease-affected families and estimates derived from populations in which unaffected individuals greatly outnumber affected individuals. Studies have explored background polygenic risk, molecular consequence of the specific variants, and biobank population characteristics as probable contributors to the discrepancy^3,12,13,17^, but factors such as mode of inheritance (MOI), single gene-level variation, and gene-disease relationship validity^19,20^ have not yet been explored.

The purpose of this genetic epidemiology study was to determine if the PGVs known to cause epilepsy in children were associated with adult-onset epilepsy in Veterans using data from the ancestrally-diverse Veterans Health Administration (VHA) Million Veteran Program (MVP) cohort^21,22^. Our primary aim was to determine the prevalence of PGVs in Veterans and estimate the penetrance of epilepsy among carriers. We also evaluated 1) the effect of MOI and gene-disease relationship validity^19,20^ on prevalence and penetrance; 2) the relative prevalence and penetrance estimates in the MVP Veteran cohort compared to previous findings by Stevelink et al. in the UKB/GEL cohorts^3^; and 3) single gene-level effects on prevalence and penetrance.

## Methods

### MVP Study Cohort, Study Period, and Whole Genome Sequencing (WGS) DNA Sequencing Data

This study was performed within the MVP cohort and uses data from the VHA electronic health record (EHR). MVP is a national research program that explores ways in which genes, lifestyle, military experiences, and exposures affect health and wellness in Veterans^21,22^. Enrollment began in 2012 and is ongoing. EHR data are available for all participants who routinely received healthcare within the VHA system between 2002 and the administrative censoring date, September 30, 2024. All MVP participants provided informed written consent. This study was approved under VHA IRB#1823793 (MVP107). STROBE (STrengthening the Reporting of OBservational studies in Epidemiology) guidelines were followed.

Of the 1,016,584 U.S. Veterans enrolled in MVP^21,22^, 102,624 (10.1%) had WGS data that could be analyzed [**Table 1**]. The primary analyses were performed within this WGS cohort. Approximately half of the WGS cohort was randomly selected, while the other half was enriched for age greater than 80 years, cardiometabolic conditions, and post-traumatic stress disorder (PTSD)^23^. WGS was performed by Illumina with mean read depth of 28x [**Supplement S1**]^23^.

**Table 1:** MVP Participant characteristics.

|  | **MVP cohort**<br>**withOUT WGS** | **MVP WGS cohort** | **Epilepsy**<br>**withOUT WGS** | **Epilepsy with**<br>**WGS** |
| --- | --- | --- | --- | --- |
| **Total (n)** | **913,919** | **102,624** | **44,491** | **6,459** |
| Age (enrollment), yrs: mean (sd) | 60.8 (14.6) | 65.6 (13.0) | 60.0 (13.0) | 62.5 (11.8) |
| range | (18 – 109) | (20 – 101) | (19 – 98) | (20 – 96) |
| Age (death/censor), yrs: mean (sd) | 67.6 (14.4) | 74.6 (12.3) | 67.4 (12.7) | 71.1 (11.3) |
| range | (21-114) | (26 – 108) | (22-105) | (27 – 102) |
| Female: n (%) | 100,410 (11.0) | 6,235 (6.1) | 5,491 (12.3) | 520 (8.1) |
| GIA: n (%) |  |  |  |  |
| AFR | 99,478 (17.9) | 23,757 (23.1) | 6,026 (19.9) | 1,694 (26.2) |
| AMR | 54,682 (9.8) | 6,383 (6.2) | 2,746 (9.0) | 349 (5.4) |
| EAS | 6,490 (1.2) | 582 (0.6) | 127 (0.4) | 13 (0.2) |
| EUR | 386,575 (69.6) | 70,812 (69.0) | 21,066 (69.4) | 4,334 (67.1) |
| SAS | 541 (0.1) | 36 (<0.1) | 25 (0.1) | 4 (0.1) |
| Other | 7,702 (1.4) | 1,054 (1.0) | 353 (1.2) | 65 (1.0) |
| Not reported | 358,451 | 0 | 14,148 | 0 |
| Self-reported race: n (%) |  |  |  |  |
| American Indian/Alaska Native | 7,623 (0.8) | 491 (0.5) | 411 (0.9) | 36 (0.6) |
| Asian | 12,531 (1.4) | 604 (0.6) | 242 (0.5) | 23 (0.4) |
| Black/African American | 159,346 (17.4) | 23,768 (23.2) | 8,846 (19.9) | 1,696 (26.3) |
| Native Hawaiian/Pacific Islander | 8,209 (0.9) | 586 (0.6) | 338 (0.8) | 39 (0.6) |
| White | 705,953 (77.2) | 76,368 (74.4) | 34,088 (76.6) | 4,636 (71.8) |
| Unknown | 20,257 (2.2) | 807 (0.8) | 566 (1.3) | 29 (0.4) |
| Self-reported ethnicity: n (%) |  |  |  |  |
| Hispanic | 68,271 (7.5) | 4,658 (4.5) | 2,929 (6.6) | 235 (3.6) |
| Non-Hispanic | 834,272 (91.3) | 97,304 (94.8) | 41,193 (92.6) | 6,190 (95.8) |
| Not reported | 11,376 (1.2) | 662 (0.6) | 369 (0.8) | 34 (0.5) |
| **Epilepsy: n (%)** | **44,491 (4.9)** | **6,459 (6.3)** | **44,491 (100)** | **6,459 (100)** |
| TBI history, any: n (%) | 156,590 (17.1) | 20,902 (20.4) | 20,198 (45.4) | 2,948 (45.6) |
| Stroke diagnosis: n (%) | 210,846 (23.1) | 35,687 (34.8) | 23,803 (53.5) | 4,079 (63.2) |
| Epi young onset (<40 years): n (%) | 5,599 (0.6) | 347 (0.3) | 5,599 (12.6) | 347 (5.4) |
| Deceased (as of 9/30/24): n (%) | 197,587 (21.6) | 43,780 (42.7) | 13,669 (30.7) | 3,016 (46.7) |

### Exposure and outcomes

The exposure was ≥1 epilepsy-associated rare variant, and outcome was an epilepsy diagnosis.

### MVP Phenotype Definitions

An established VHA-validated algorithm^6,24,25^ identified epilepsy cases from VHA EHR data. The algorithm requires a combination of ≥2 diagnosis codes for epilepsy and/or convulsions on two separate dates, as well as concurrent anti-seizure medication (ASM) prescriptions (excluding gabapentin/pregabalin) with a minimum of 3 filled prescriptions from VHA pharmacies^6,24,25^. MVP participants were deemed epilepsy cases if they met algorithm criteria for epilepsy at any time between 2002 and censorship or death. Cases were classified as “young onset” epilepsy if diagnostic codes were found in EHR prior to age 40. TBI history and stroke history were determined by previously validated algorithms using EHR data^9,26^.

### Comparator Cohorts – UKB and GEL

Stevelink et al. analyzed 42,863 individuals from the GEL cohort and 392,123 individuals from UKB who had WGS data available for rare variant analysis^3^. The GEL cohort includes 78,000 patients with rare diseases, cancer, and unaffected relatives recruited via centers across the UK with a mean age of 30 years (range 0 – 99). The UKB cohort includes 500,000 individuals aged 40–69 years at recruitment. People with profound intellectual disability were likely excluded because capacity to provide informed consent was required.

### Epilepsy Gene and Variant Identification and Classification

To determine gene-disease validity^19,27^, we used ClinGen’s “Epilepsy Gene Expert Curation Panel” website^20^ (accessed 7/25/2025) to identify 119 evidence-based expert-curated epilepsy candidate genes. ClinGen labels each gene by associated epilepsy type (e.g., DEE, temporal lobe epilepsy), MOI, and “classification” or level of evidence as an epilepsy gene (i.e., “definitive”, “strong”, “moderate”, “limited”, “disputed”, and “refuted”) [**Supplement S2**]. When more than one classification was provided, our analyses defaulted to the higher level of classification. For example, definitive/moderate genes were assigned to definitive.

We then utilized the ClinVar^28^ website (accessed 11/10/25) to download all pathogenic and/or likely pathogenic (P/LP) single nucleotide variants (SNVs) reported and classified within the 119 genes. *USP25, CHRNA7, CRH*, *EFHC1* were excluded because no PGVs were reported on ClinVar, and *SIK1* was excluded because it could not be queried in MVP due to technical limitations and WGS locus quality. An additional 51 AD genes not yet classified by ClinGen but reported in Genes4Epilepsy^10^ were analyzed to compare to the UKB/GEL study^3^. Thus, a total of 165 epilepsy gene candidates were evaluated in the MVP WGS cohort. No manual curation of variants was performed. Variants classified as a “variant of uncertain significance (VUS)” or having “conflicting evidence” were not included in the analysis.

A list of all ultrarare variants queried by Stevelink et al^3^ (n=4,294 variants across 92 AD-labeled DEE genes) was provided for this analysis. In contrast to our MVP study, the UKB/GEL study excluded PGVs in epilepsy genes that were not specifically associated with an epilepsy phenotype in ClinVar [**Supplement S3**].

### Statistical analysis

We restricted the primary analyses of epilepsy prevalence and penetrance to ultrarare PGVs (VAF<0.0001) in epilepsy candidate genes, as done in Stevelink et al^3^. We conducted sensitivity analyses that included rare PGVs (VAF<0.01). Epilepsy status was compared between carriers (individuals carrying ≥1 PGV) and non-carriers, and by carriers of one classification of PGV and another (i.e., definitive and limited/disputed/refuted), by odds ratios (OR) and 95% Wald confidence intervals (CIs). CIs excluding 1 were considered statistically significant. When we compared the MVP and UKB/GEL cohorts, we restricted our analyses of epilepsy prevalence and penetrance to the 91 AD-labeled genes queried within all three cohorts. Statistical analyses were performed using R version 4.3.2.

To calculate penetrance, we assessed the proportion of individuals with epilepsy diagnoses among Veteran carriers. We corrected for ascertainment bias in penetrance estimates using Bayes’ theorem as previously described^3^ [**Supplement S4**]. The UKB/GEL assumed a population prevalence of 0.7%^2–4,29^ for childhood-onset epilepsy. For MVP, we used the population prevalence of the full MVP cohort (5.0%) to calculate penetrance.

### Data availability

The datasets used in this study are not publicly available due to privacy concerns. Patient-level data are available to all VA researchers with appropriate IRB approvals. PGV summary statistics will be submitted to the NIH Database of Genotypes and Phenotypes (dbGaP).

## Results

### MVP Epilepsy Prevalence and Penetrance

Among the 102,624 MVP participants with available WGS data, epilepsy prevalence was 6.3% (n= 6,459); 0.3% (n=347) had epilepsy diagnosis before age 40 [**Table 1**]. Epilepsy prevalence for the full MVP cohort was 5.0% (n=50,956/1,016,584). Within the WGS cohort, mean age at study enrollment was 65.6 years (standard deviation [SD] 13.0 yrs, range 20-101), mean age at censorship or death was 74.6 (SD 12.3, range 26-108), and 6.1% (n=6,237) were female. Genetically inferred ancestry (GIA) was 23.1% African (AFR; n=23,757), 6.2% Admixed American (AMR; n=6,383), 0.6% East Asian (EAS; n=582), 69.0% European (EUR; n=70,812), <0.1% South Asian (SAS; n=36), and 1.0% “Other” (n=1,054).

Stroke and TBI history were more common in participants with epilepsy (63.2% and 45.4%, respectively) than within the full cohort (34.8% and 20.4%).

#### Approximately 1% of MVP participants carried ultrarare PGVs in epilepsy genes

Among the MVP WGS cohort, 1,041 participants (1.0%) were carriers of ≥1 unique ultrarare PGV within 94 of the 147 unrefuted/undisputed/limited epilepsy-associated genes queried **[Supplementary Table ST1 G ST2]**. Most ultrarare carriers had only 1 PGV, while 11 participants carried 2 PGVs (only 1 of whom had epilepsy).

#### Refuted, disputed and/or limited genes were not associated with epilepsy

Carriers of ultrarare PGVs in genes classified by ClinGen as definitive had greater than three times the odds of epilepsy diagnosis than carriers of PGVs in genes classified as limited, refuted, and/or disputed (OR 3.22 [1.03–16.26]). Carriers of limited/refuted/disputed PGVs did not have increased odds of epilepsy diagnosis compared to non-carriers (OR 0.42 [0.13–1.31]) **[Supplementary Table ST1]**, justifying exclusion of these variants from further analysis. No carriers of limited or disputed genes had epilepsy. The highest epilepsy prevalence in carriers of a single refuted gene was 4.6% (*CLCN2*), which fell below the WGS cohort prevalence of 6.3% **[Supplementary Table ST2]**.

#### Penetrance of AD-labeled ultrarare PGVs ranged between 7.5–12.8% depending on gene grouping

Carriers of PGVs found in the 97 unrefuted/undisputed AD-labeled genes had an epilepsy prevalence of 10.3% and increased odds of epilepsy compared to non-carriers (OR 1.72 [1.09–2.70]); however, penetrance was low at 8.3% **[Figure 1, Supplementary Table ST1]**. When we restricted analyses to PGVs in the 40 “definitive” AD genes, the effect size was no longer statistically significant (OR 1.54 [0.76–3.06]), prevalence was 9.4%, and the penetrance estimate was 7.5%. Conversely, when we restricted analyses to the 51 ClinGen-unclassified additional DEE genes included in the UKB/GEL study, the effect size was greater (OR 2.79 [1.37–16.64]), prevalence was 15.8%, and the penetrance estimate was 12.8%. Carriers of PGVs in the 6 “moderate” AD genes did not have increased odds of epilepsy (OR 0.95 [0.30–3.05).

**Figure 1:**
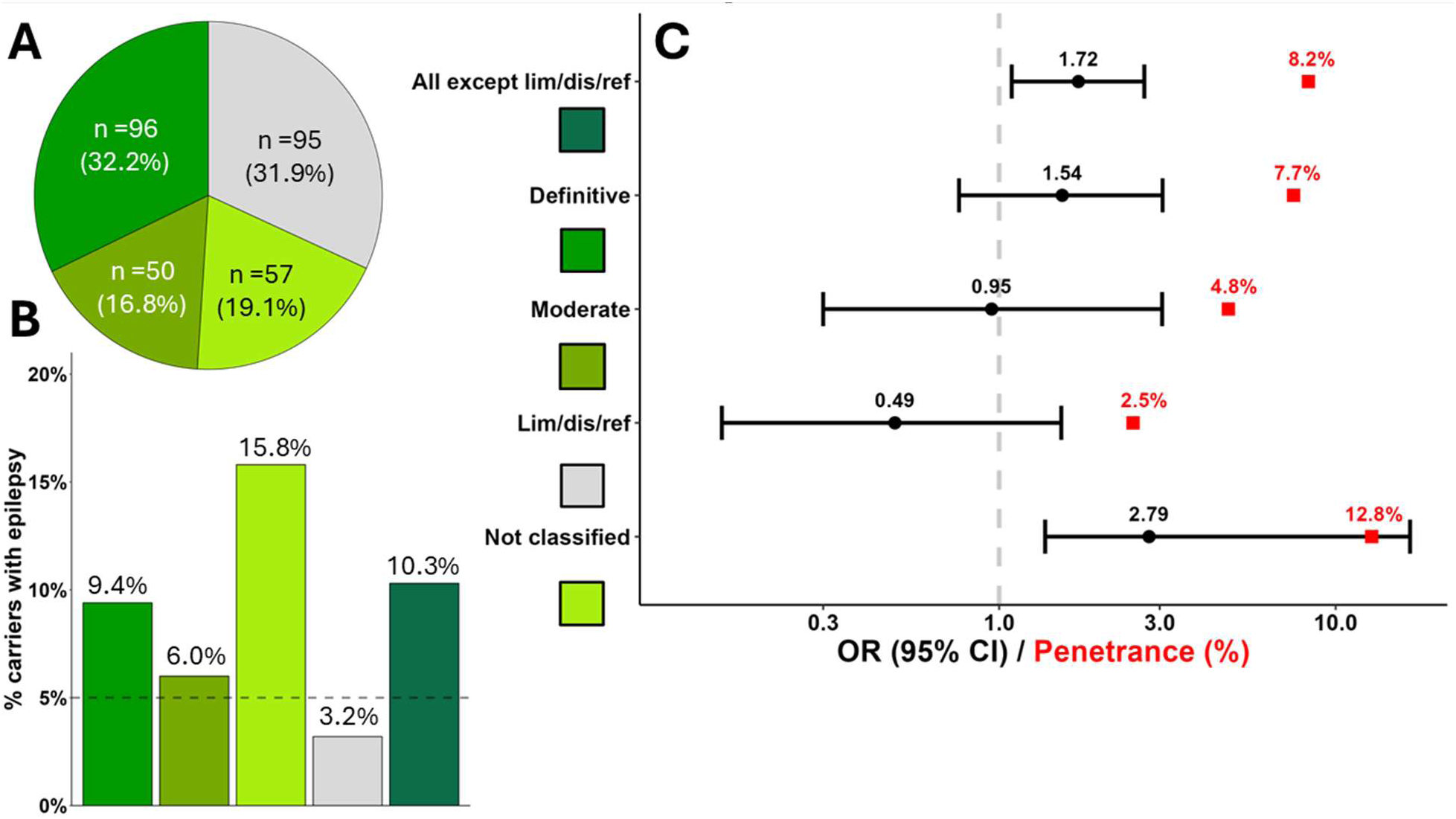
Prevalence, Odds and Penetrance of Epilepsy in Carriers of AD-labeled PGVs by ClinGen Classification. A) Carriers of AD-labeled PGVs, colored by ClinGen classification. B) Prevalence (%) of epilepsy among carriers by ClinGen classification; the dashed line marks epilepsy prevalence in the full MVP cohort (5%). C) Odds of epilepsy among carriers (OR = black dots) with Wald 95% CI (black whiskers) and penetrance estimates (red squares) for each ClinGen classification, presented on the log scale. Abbreviations: AD, autosomal dominant; CI, confidence interval; OR, odds ratio; MVP, Million Veteran Program; PGV, germline pathogenic variant

#### Carriers of autosomal recessive ultrarare PGVs had increased odds of epilepsy diagnosis

Carriers of PGVs in the 38 autosomal recessive (AR)-labeled genes (excluding 3 limited genes) had an epilepsy prevalence of 8.8%, a penetrance estimate of 7.1%, and increased odds of epilepsy compared to non-carriers (OR 1.45 [1.13–1.85]) [**Figure 2**]. The 19 carriers of PGVs in the 6 undisputed/unrefuted AD/AR-labeled genes did not have increased odds of epilepsy (OR 0.83 [0.11–6.20]) and were not incorporated into the AR or AD analyses due to ambiguity [**Figure 3**]. There was no significant difference in epilepsy prevalence between AD and AR PGV carriers (OR 1.19 [0.71–1.99]).

**Figure 2:**
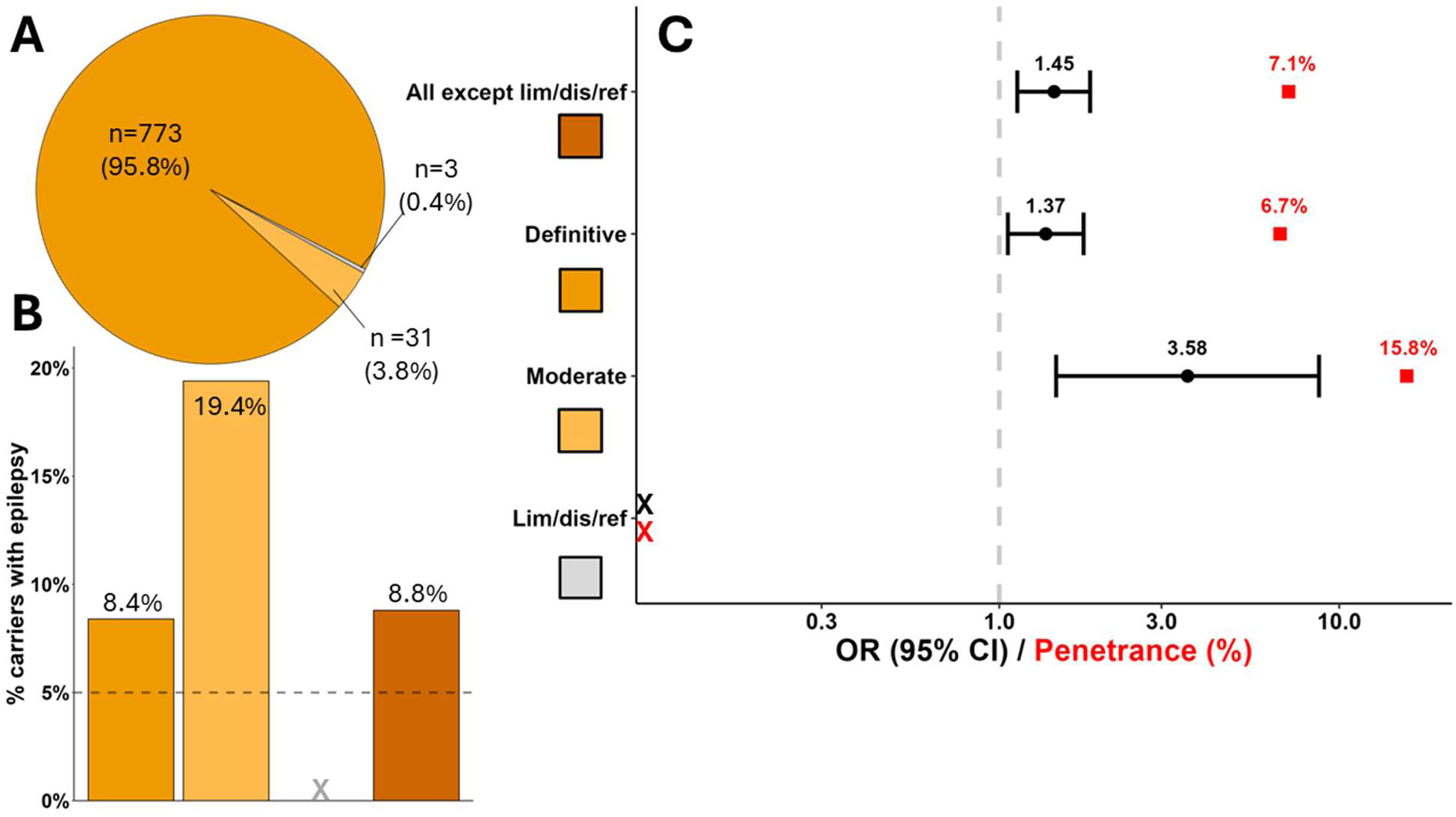
Prevalence, Odds and Penetrance of Epilepsy in Carriers of AR-labeled PGVs by ClinGen Classification. The pie chart depicts all carriers of AR-labeled PGVs divided/colored by ClinGen classification. The bar chart illustrates the prevalence (%) of epilepsy among carriers by ClinGen classification and the dashed line marks epilepsy prevalence in the full MVP cohort (5%). The forest plot represents the odds of epilepsy (OR = black dots) with Wald 95% CI (black whiskers) with the calculated penetrance estimates (red squares) for each ClinGen classification. Carriers of AR-labeled PGVs classified as definitive (n=773) or moderate (n=31) had significantly increased odds of epilepsy. No carriers of PGVs classified as limited (n=3) had epilepsy (indicated by X on plots B and C). Abbreviations: AR, autosomal recessive; CI, confidence interval; OR, odds ratio; MVP, Million Veteran Program; PGV, germline pathogenic variant

**Figure 3:**
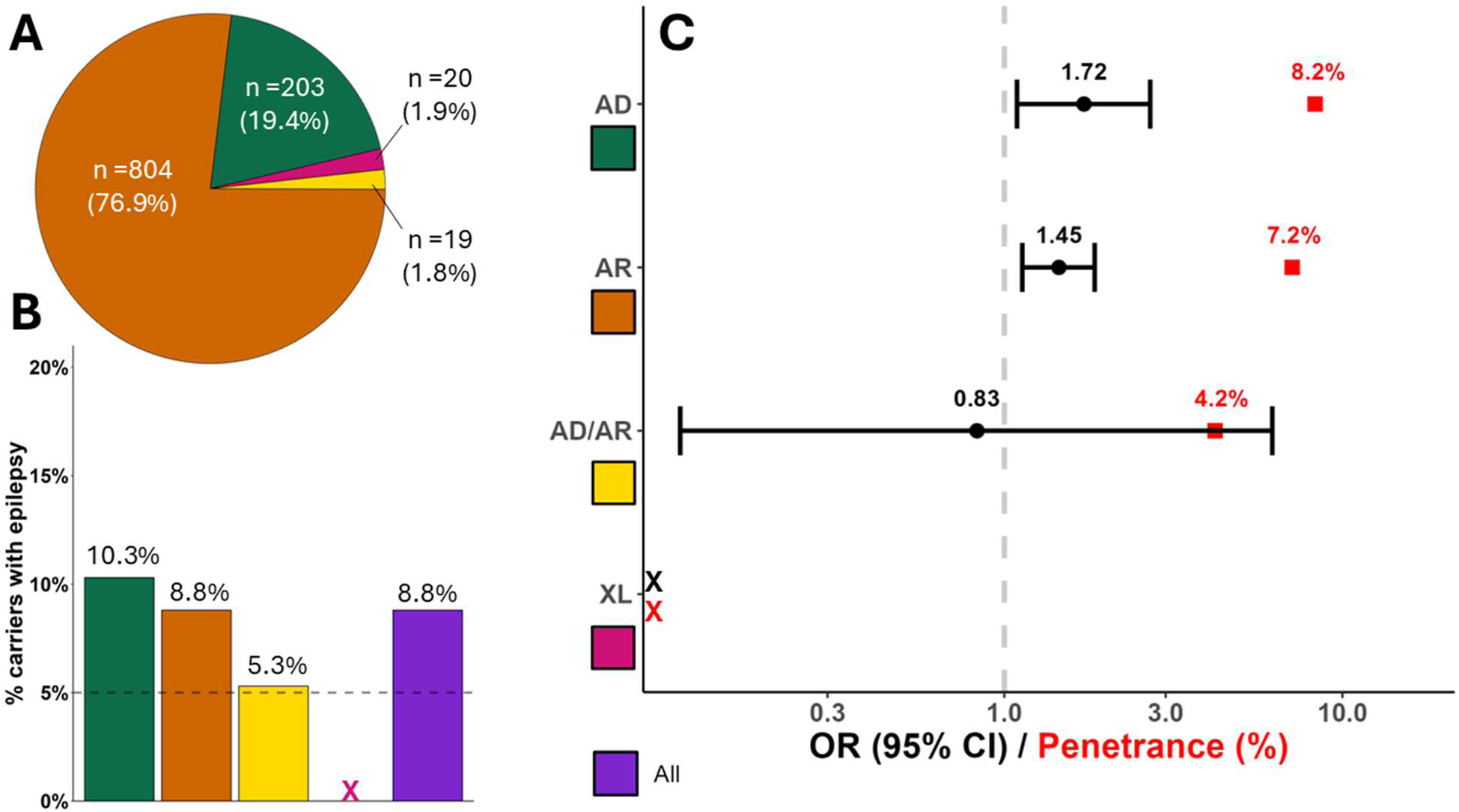
Prevalence, Odds and Penetrance of Epilepsy in Ultrarare PGV Carriers by MOI. The pie chart depicts PGV carriers divided/colored by labeled MOI (excluding limited, refuted or disputed genes). The bar chart illustrates the prevalence (%) of epilepsy among carriers by MOI and the dashed line marks epilepsy prevalence in the full MVP cohort (5%). The forest plot represents the odds of epilepsy (OR = black dots) with Wald 95% CI (black whiskers) with the calculated penetrance estimates (red squares) for each MOI. No carriers of XL PGVs (n=20) had epilepsy (indicated by X on plots B and C). Abbreviations: AD, autosomal dominant; AR, autosomal recessive; CI, confidence interval; OR, odds ratio; MOI, mode of inheritance; MVP, Million Veteran Program; PGV, germline pathogenic variant

#### No carriers of X-linked ultrarare PGVs had epilepsy in our MVP sample

Only 20 individuals carried PGVs in 4 of the 6 unrefuted/undisputed X-linked (XL) genes queried, and none had epilepsy. All 20 carriers were male; 13 were carriers for 9 unique PGVs in *PCDH19* [**Supplementary Tables ST1 G ST2**], a definitive epilepsy gene for which the epilepsy phenotype predominantly manifests in heterozygous females^30^.

#### Rare-only PGVs were not associated with increased odds of epilepsy in carriers

Rare PGVs were found in 1.9% (n=1,955) of the WGS cohort [**Supplementary Table ST3**]. Of the 9,468 unique ClinVar-classified PGVs in epilepsy genes queried, 95.6% (n=517/541) of PGVs detected were ultrarare and 24 were rare-only (0.0001<VAF<0.01).

Most rare-only PGVs (n=22/24) were found in <90 individuals each (0.0001<VAF<0.001). A *TTP1* splice acceptor SNV (rs56144125) was the most common PGV found in 167 individuals (9.0% had epilepsy). Most rare PGV carriers carried only 1 PGV, but 23 individuals carried 2 (in different genes) and only 1 had epilepsy. Carriers of rare PGVs did not have significantly increased odds of epilepsy diagnosis compared to non-carriers (AD-labeled: OR 1.56 [0.99–2.44]; AR-labeled: OR 1.18 [0.98–1.42]).

### Comparison of MVP, UKB and GEL Cohort Prevalence and Penetrance

#### Only 2 unique ultrarare PGVs were detected in all 3 biobank cohorts

Among the 91 AD epilepsy genes analyzed in all 3 cohorts, only 2 of the 2,982 shared PGVs queried were found in all 3 biobanks. A *CHD2* LP missense SNV (rs150951454, gnomAD VAF 0.00001) was found among 43 carriers (MVP 3, UKB 37, and GEL 3), only 1 of whom had epilepsy (2.3%). A *SCN1A* P/LP missense SNV (rs121918782, gnomAD VAF 0.00001) was found among 10 carriers (MVP 3, UKB 6, GEL1), none of whom had epilepsy.

#### Penetrance estimates are similar across cohorts, but odds of epilepsy are divergent

When we calculated penetrance using the identical 2,982 ultrarare SNVs queried in all 3 cohorts, the MVP penetrance estimate of 8.8% falls between UKB’s 12.4% and GEL’s 6.2% [**Figure 4a-c, Supplementary Table ST3**]. We find similar carrier frequencies across cohorts (MVP 0.06%, UKB 0.06%, GEL 0.08%), and VAF in carriers without epilepsy is identical (0.06%); however, VAF in individuals with epilepsy (MVP 0.11%, UKB 1.1%, GEL 0.54%) and epilepsy prevalence in carriers (MVP 10.9%, UKB 3.9%, and GEL 29.4%) differ. Consequently, the odds of epilepsy in carriers compared to non-carriers were significantly increased in UKB (OR 20.24 [10.35–39.56]) and GEL (OR 9.34 [4.46–19.56]) but had a weaker effect size in MVP (OR 1.83 [0.86–4.01]). When we did *not* restrict the analysis to identical PGVs queried across cohorts, OR effect sizes were minimally affected, but the results became statistically significant for MVP (OR 1.72 [1.04–2.85]) [**Figure 4d-f**]. Thus, exclusion of non-epilepsy specific PGVs reduced sample size and did not meaningfully change effect size.

**Figure 4:**
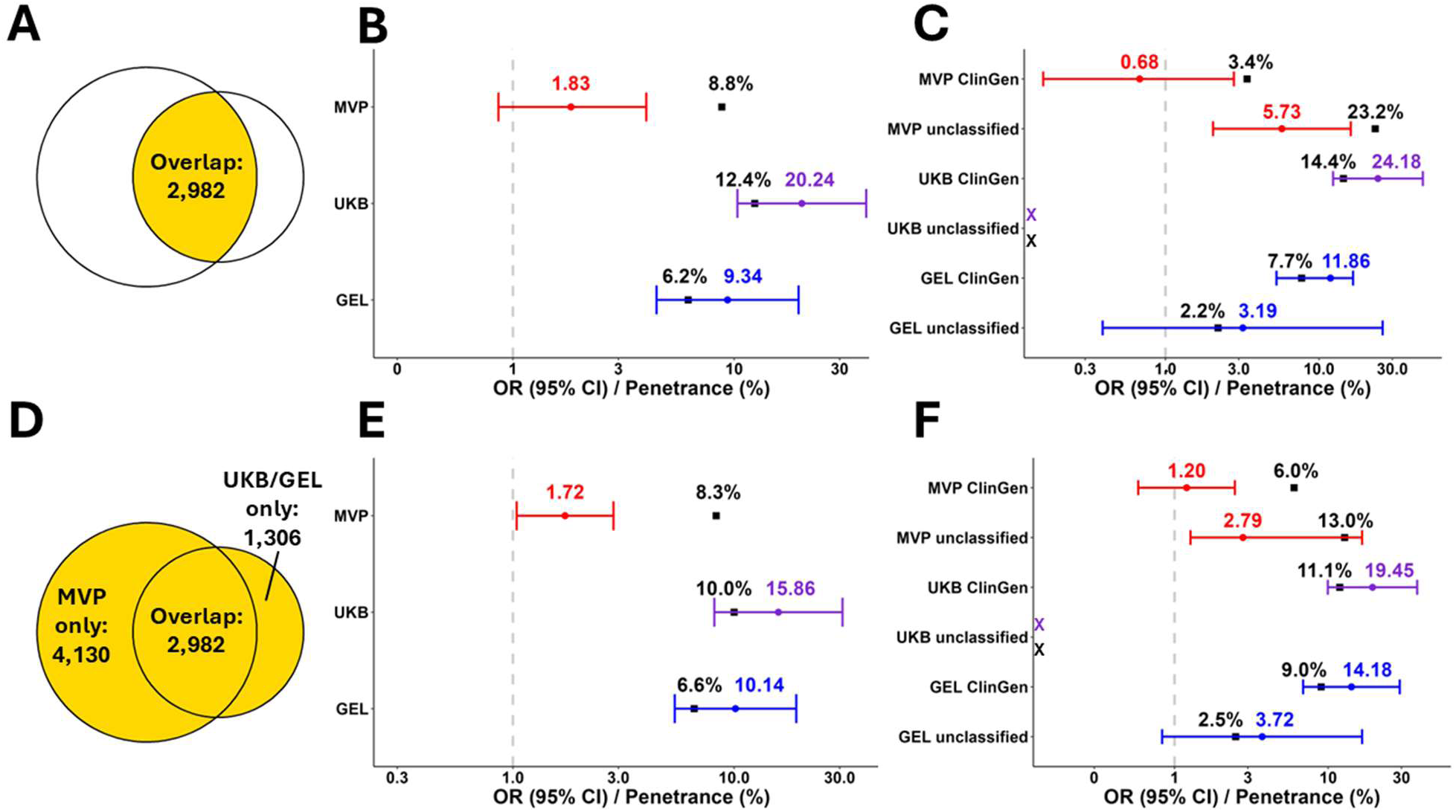
Comparison of Odds and Penetrance of Epilepsy in Ultrarare PGV Carriers Across Cohorts. To compare the odds and penetrance of epilepsy across cohorts for the 91 AD-labeled genes, we separately analyzed “overlapping” PGVs (a–c) that were queried identically across cohorts (n=2,982), versus “expanded” PGVs (d–f) that each study defined somewhat differently (MVP n=7,112; UKB/GEL n=4,288). MVP PGVs were restricted to SNVs labeled as pathogenic or likely pathogenic by ClinVar and no manual curation was performed (i.e., no filtering of epilepsy-specific PGVs). UKB/GEL PGVs included indels and the authors had performed manual curation of variants, excluding PGVs that were not explicitly associated with epilepsy or DEE in ClinVar. The Venn diagrams (a, d) demonstrate the numbers of unique PGVs queried (yellow) for the overlapping (a-c) versus expanded (d– f) analyses. The forest plots (b, e) represent the odds of epilepsy (OR = colored circles) with Wald 95% CI (colored whiskers) with the calculated penetrance estimates (black squares) for all 91 genes with overlapping PGVs (b) and expanded PGVs (e). Forest plots (c, f) compare the odds of epilepsy and penetrance when the 91 genes are divided into the 40 ClinGen-classified (definitive or moderate) “ClinGen” genes and the 51 ClinGen “unclassified” genes Abbreviations: CI, confidence interval; DEE, developmental and epileptic encephalopathy; GEL, Genomics England; OR, odds ratio; MVP, Million Veteran Program; PGV, germline pathogenic variant; SNV, single nucleotide variant; UKB, UK Biobank

### Epilepsy Gene-Level Ultrarare PGV Descriptive Analyses

#### In MVP, the top 20 genes associated with higher epilepsy prevalence included 7 AR-labeled genes and 1 AR/AD-labeled gene

Among the 165 epilepsy genes queried in MVP, 46 genes (27.9%) had at least 1 PGV carrier with epilepsy [**Figure 5**], 57 (34.5%) had no carriers with epilepsy, and 62 (37.6%) were undetected (i.e., no carriers) [***Supplementary Table ST4***]. Four genes had 100% penetrance in MVP (*LGI1, ATP1A3*, *KCNA3* and *UBE3A);* however, penetrance should be interpreted with caution because only 1 or 2 participants carried PGVs in each gene.

**Figure 5:**
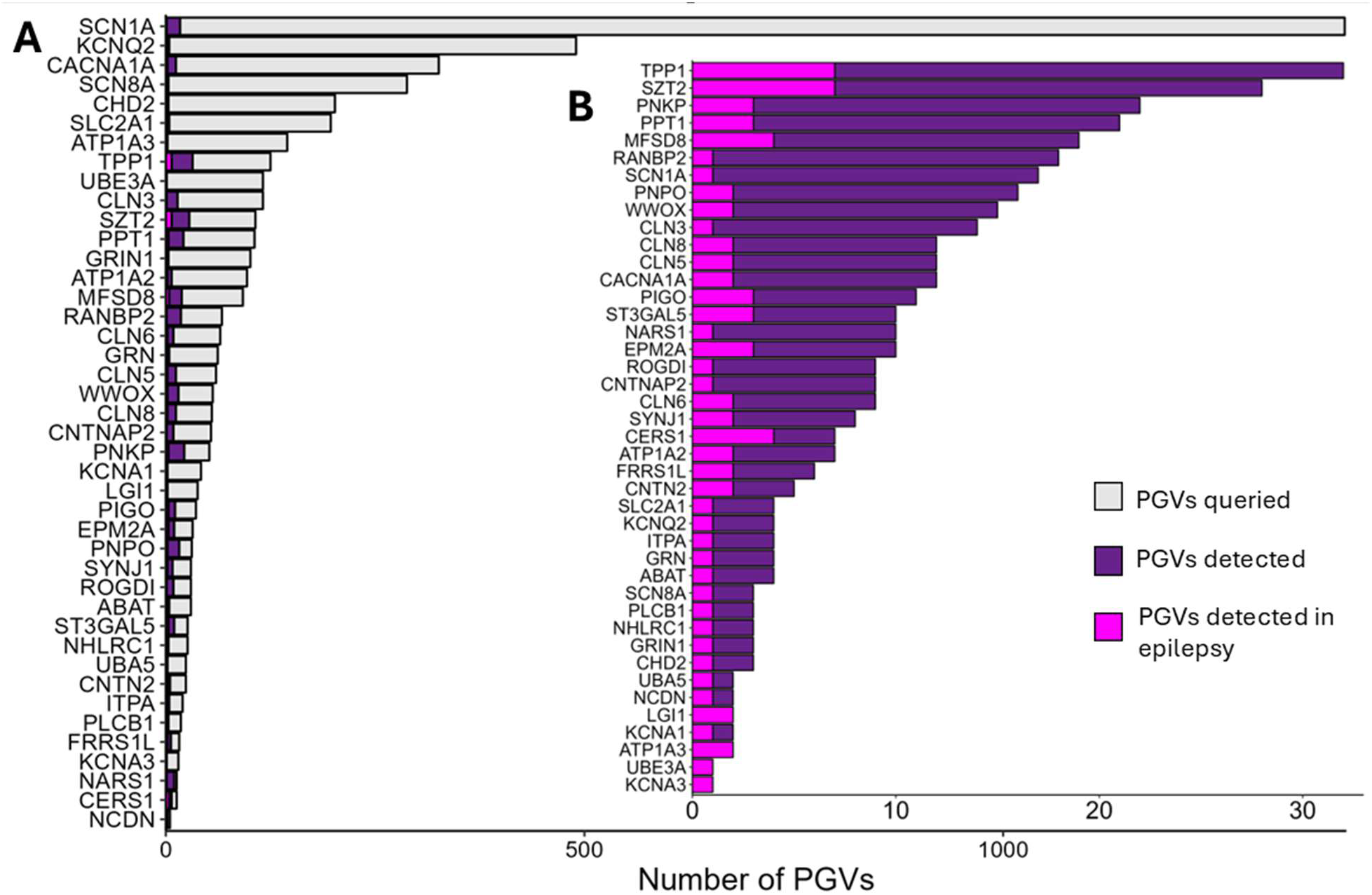
MVP-based gene-level comparisons of epilepsy genes in which at least 1 ultrarare PGV carrier had epilepsy. A) All unique PGVs **queried** per gene. B) All unique gene-associated PGVs **detected** in MVP. C) Ratio of gene-associated PGVs detected *with* epilepsy cases (magenta) to those *without* epilepsy cases (purple). Abbreviations: MVP, Million Veteran Program; PGV, germline pathogenic variant

Among the top 20 genes for epilepsy prevalence (range 13.16–100%), ClinGen classified most (n=13) as definitive, except for *ABAT* (moderate) and 6 unclassified genes. ClinGen’s MOI designation was AR for 7 genes, AD for 6, AD/AR for 1, and unclassified for 6 (presumed AD by Stevelink et al^3^ and Genes4Epilepsy^10^). Most genes (n=15) had ≤5 carriers, precluding significance testing due to small sample size. Two of the 3 genes with 12 or more PGV carriers were labeled AR (*PIGO* n=5/38 cases/carriers; *ST3GAL5* n=4/21) and 1 was unclassified (*CACNA1A* n=2/12).

#### Among AD-labeled genes queried in all 3 cohorts, ultrarare PGVs and epilepsy cases were more commonly found in certain genes and undetected in others

Among the 91 AD genes queried, 12 genes (13.2%) had ≥1 ultrarare PGV detected in each of the cohorts and 35 genes (38.5%) had zero PGVs detected [**Figure 6, Supplementary Table ST5]**. 21 genes (23.1%) had ≥1 case of epilepsy diagnosed amongst ultrarare PGV carriers and 12 genes had >10 carriers. Amongst ultrarare PGV carriers, 35 genes (38.5%) had zero epilepsy cases. The 12 genes with the greatest number of PGVs detected were also genes in which a greater average number of PGVs were queried: 295 PGVs per gene (n=3,420/12) versus 78 PGVs per gene for the rest (n=7,113/91). For example, only 17 unique variants were found in the *SCN1A* gene amongst 1,408 queried.

**Figure 6:**
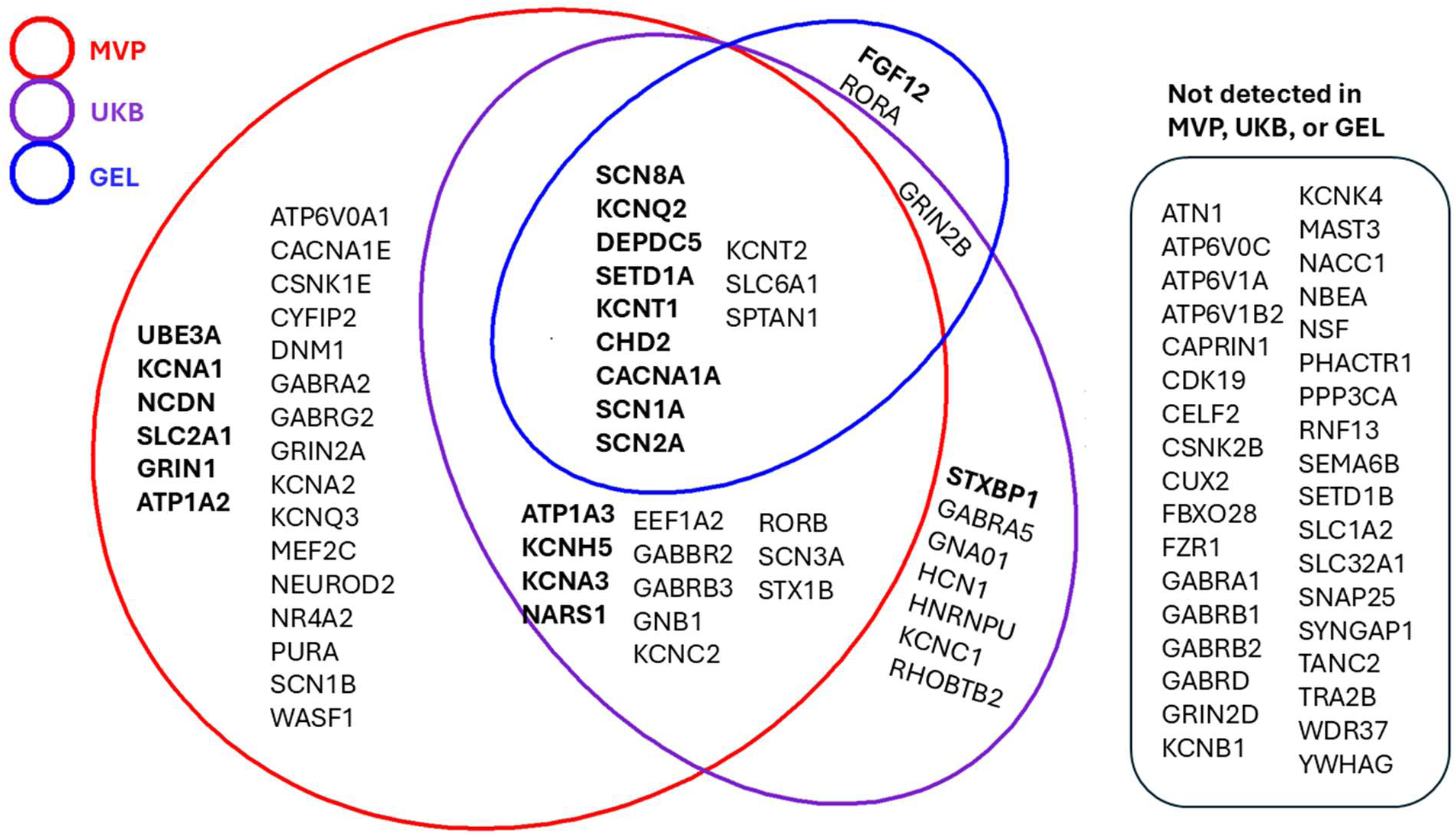
Venn diagram of epilepsy genes in which PGVs were detected among the 3 different biobanks: Bold font indicates that at least 1 PGV carrier had epilepsy and normal font indicates no carriers with epilepsy. Abbreviations: PGV, germline pathogenic variant

Among the 35 genes with zero detected PGVs, most (n=18) had <10 PGVs queried; however, 7 had >40 PGVs queried: *SYNGAP1*, *SETD1B, KCNB1*, *CSNK2B*, *GABRA1, NBEA*, and *GABRB2*. Among the 35 genes with no epilepsy cases, the majority (n=31) had ≤8 carriers found across cohorts, but 4 genes with no cases had ≥14 carriers each: *PURA*, *SPTAN1, KCNT2*, and *SCN1B*.

## Discussion

Approximately 1% of Veterans with WGS in MVP carried ultrarare PGVs in epilepsy-associated genes. Most carriers (∼90%) had not been diagnosed with epilepsy, though the odds of epilepsy in carriers were 1.45 and 1.72 times higher in carriers of AD and AR PGVs, respectively, than in non-carriers in MVP. Penetrance estimates were low (8.2% and 7.1% for AD and AR PGVs, respectively), yet within range of the UKB and GEL estimates. Carriers of definitive epilepsy PGVs were over 3 times more likely to be diagnosed with epilepsy than carriers of limited/refuted/disputed genes, which is comparable to previous estimates of the magnitude of familial risk^31^ and supports ClinGen’s evidence-based expert consensus assessments^19,27^. However, AD-labeled PGV carriers were not significantly more likely than AR-labeled PGV carriers to be diagnosed with epilepsy. Moreover, AR PGV carriers had 45% increased odds of developing epilepsy than non-carriers.

Based on these findings, if clinical WGS or epilepsy multigene panels were performed in a similar population, approximately 1 of 100 would return pathogenic results, and approximately 1 in 5 PGVs detected would be in AR-labeled genes and therefore considered incidental or indicative of carrier status. These counterintuitive numbers can be partially explained by the unique characteristics of the U.S. Veteran population^21,22^, potentially lower penetrance or variable expressivity of the detected variants^32^, and population genetics factors^33–35^. The MVP population is enriched for adult-onset disease because people with active epilepsy and other disabling childhood-onset conditions are ineligible for military enlistment. Thus, Veterans are unlikely to harbor highly penetrant childhood-onset monogenic disease beyond self-resolving childhood epilepsies or febrile seizures (which would not be captured in the EHR). Additionally, MVP is a more ancestrally diverse cohort than UKB and GEL, so there are likely more undiscovered PGVs, particularly in participants of AFR, AMR or “other” ancestry who comprise 31% of the MVP WGS cohort; the different genetic backgrounds may alter a PGV’s phenotypic effect. Finally, MVP is male-predominant and thus we likely see lower penetrance estimates for PGVs whose phenotypes manifest more frequently in females^36,37^. For example, none of the 13 *PCDH1S* PGV carriers were diagnosed with epilepsy, which typically only manifests in heterozygous females^30^.

The most surprising findings in our study were that estimated epilepsy phenotype prevalence and penetrance in our sample did *not* significantly differ between carriers of AD- and AR-labeled PGVs, and that AR-labeled PGV carriers were at increased odds of epilepsy compared to non-carriers. According to classically defined genetic “dominance,” one dominant PGV is sufficient to increase disease risk whereas two recessive PGVs are required to substantially increase risk^33^. Possible explanations for our findings include specific features of our sample, complex gene-gene or gene-environment interactions, or mislabeling of a gene or variant’s dominance. The MVP sample excluded people with more severe childhood onset epilepsies, so homozygous individuals were excluded due to population selection bias. Of the 11 participants in MVP with 2 ultrarare PGVs, only one had epilepsy, so our findings cannot be explained by complex heterozygotes carrying 2 known PGVs. VUS, conflicting evidence variants, and PGVs yet to be discovered were not analyzed in this study, offering one potential (currently unquantifiable) reason for the elevated epilepsy risk in AR PGV carriers. Genetic population structure (i.e. differences in genetic background risk due to ancestry, age, or sex) may contribute to these findings^38,39^, but ancestry and age distributions were similar across AD-labeled and AR-labeled carriers and a sensitivity analysis of only males in MVP yielded similar results. One possibility is that carrying certain recessive PGVs confers a component of epilepsy risk, particularly in adults who have had a longer lifetime to accumulate somatic variants, epigenetic changes, or in certain adult-onset phenotypes like post-stroke or post-traumatic epilepsy that have not been well-represented in genetic studies to date. Our MVP sample was older and exposed to more epilepsy risk factors, particularly TBI and stroke, than most samples in epilepsy genetic studies, including the UKB/GEL samples. AR-labeled PGVs may express at higher rates in later life in monogenic carriers. Certain alleles in recessive epilepsy genes could confer a small component of the polygenic risk for late-onset epilepsy, the genetics of which require further study and could include digenic or oligogenic inheritance, age-related somatic variation, or gene-environment interactions. Finally, we used ClinGen gene-level classifications to label genes as AD or AR, but it is likely more accurate to determine dominance at a variant level^19,33^. Regardless, this surprising finding must be validated in other cohorts, and ongoing refinement of how dominance for genes and variants are determined is needed^19,32,33^.

The ∼1% prevalence estimate for epilepsy-associated ultrarare PGVs in U.S. Veterans is likely an underestimate. By restricting our analysis to 165 ClinGen expert-curated epilepsy genes^20^ and the UKB/GEL study’s genes^3^, we analyzed a minority of the >1,000 genes implicated in epileptogenesis^10^. Moreover, additional genes and non-coding variants are still being discovered as associated with the epilepsy phenotype. By restricting our MOI analyses to currently ClinVar-classified PGVs, we excluded “conflicting evidence” variants, VUS, and countless unpublished rare SNVs that may ultimately be (re)classified as pathogenic. Although some variants may be downgraded from pathogenic to benign or may have been classified as pathogenic for conditions other than epilepsy/DEE,^32,34^, we hypothesize that the proportion of PGVs currently misclassified or misattributed are far outnumbered by the PGVs yet to be discovered and/or published. Our estimates of prevalence are also hampered by lack of a consistent time period denominator across diverse longitudinal cohorts of varying ages, birth years, sexes, and ancestries^40–42^. Finally, including AR-labeled PGVs in this analysis gave us a higher cumulative prevalence of PGVs than if we had restricted our analysis to AD-labeled genes only.

Penetrance estimates were low for AD-labeled PGVs in the MVP cohort, but fell within range of those in the UKB and GEL cohorts and other similar population-based studies^3,16,43,44^, thus validating Stevelink et al’s findings^3^. However, our study illustrates how drastically penetrance estimates can vary when selecting different genes and variants or employing different population-based phenotype prevalence assumptions. Although the MVP and UKB/GEL studies used age-specific prevalence assumptions specific to their sample populations in penetrance calculations, neither study adjusted for sex or ancestry, which is recommended for optimal penetrance estimation^32,34^. The penetrance formula our studies used relies on assumptions regarding epilepsy prevalence in the population (e.g. that prevalence does *not* vary by age, epoch, sex, ancestry, etc.), which may not hold upon further study. Future studies would benefit from adjustment for these critical demographic covariates and optimal penetrance estimates would incorporate survival analysis modeling, which is limited in biobank studies due to immortal person time bias and population selection bias.

A major confounding factor when comparing the MVP and UKB/GEL results is that epilepsy prevalence in the MVP WGS (6.3%) cohort is not only higher than in the UKB (0.2%) and GEL (4.3%) study, but also higher than many other population-based estimates used to calculate penetrance (0.7-2.9%) and the full MVP cohort prevalence (5.0%). Potential reasons for this include the older age of MVP participants (particularly the WGS cohort), high prevalence of comorbidities (e.g., stroke, TBI) that increase seizure risk^7,8^, and methods of epilepsy ascertainment. A recent study of the full VHA population, a younger cohort than the MVP cohort, calculated epilepsy prevalence to be 2.9% in post-9/11 Veterans with TBI^7^. The UKB/GEL study^3^ limited epilepsy diagnosis to <18 years of age, which resulted in 0.2% epilepsy prevalence; however, roughly 5% of UKB subjects had an epilepsy ICD code at some point in their life, which approximates that of the full MVP cohort. If we had restricted our analysis to Veterans diagnosed with epilepsy before 40 years old, epilepsy prevalence would have been 0.3%; however, ascertainment was limited prior to 2002 so young-onset epilepsy is likely undercounted in MVP, particularly in older individuals. Importantly, MVP’s 6.3% represents cumulative prevalence of epilepsy during the study period, which ranges from 0.5-22 years for each Veteran depending on age and duration of VHA-based care. In contrast, the 1.1% point prevalence reported by the Center for Disease Control (CDC)^1^ represented adults who reported “active epilepsy” at the time of CDC’s 2013 survey, which is lower than a cumulative prevalence by definition. Another potential reason for the higher prevalence is that the MVP WGS subset was purposefully enriched for older Veterans, psychiatric diagnoses, and cardiometabolic conditions^23^, all of which are associated with increased risk of epilepsy; evidence for this contributive factor is that cumulative epilepsy prevalence in the full 1,016,584 MVP cohort is lower at 5.0%.

Finally, while the GEL cohort is enriched for undiagnosed and/or complex conditions, UKB is recognized to have a healthier cohort^45^ than MVP’s VHA cohort^22^, since VHA cares for more Veterans with chronic and/or disabling conditions than Veterans who utilize employer-based private healthcare insurance outside VHA.

Emerging biobank literature offers numerous explanations for the low penetrance of AD-inherited epilepsy genes. A recent population-based study of predicted loss of function (pLoF) variants in the Genome Aggregation Database (gnomAD) found that “rescue” (i.e., modifying) variants explained some of the incomplete penetrance for 77 haploinsufficient genes associated with severe, early onset disorders, including numerous epilepsy genes detected in MVP: *CHD2*, *GRIN2B*, *SCN1A*, *SCN2A*, *SLC2A1,* and *STXBP1*^16^. The study also noted genetic ancestry group-specific incomplete penetrance, which was related to certain ancestries carrying pLoF and rescue variants in cis^16^. Moreover, the gnomAD study observed a trend of higher age distribution in a large subset of pLOF carriers whose incomplete penetrance was unexplained, suggesting somatic origin (i.e., somatic variants resembling germline variants due to clonal expansion)^16^. Other possibilities include variable expressivity, laboratory error (i.e., false positive detections in the WGS pipeline due to sequencing artifact), ClinVar misclassification leading to high rates of “false positive” PGV labels (described in other studies^32^), and inclusion of variants associated with milder and/or alternative phenotypes (e.g., migraine).

Our findings suggest that the PGVs detected in the MVP and UKB populations may be less penetrant and less deleterious to protein function than variants that typically cause DEE. We saw much higher prevalence of epilepsy in PGV carriers in the GEL cohort than MVP and UKB, likely because GEL was enriched for rare and earlier onset disease.

Additionally, we found no PGVs in 35 of the epilepsy genes analyzed in all 3 cohorts, several of which are recognized to be highly intolerant to variation^46,47^, such as *SYNGAP1*, *GABRA1,* and *KCNB1*. On the other hand, we know variants in *SCN1A* can be asymptomatic or associated with other mild non-epilepsy phenotypes like migraine or isolated febrile seizures^48,49^, so finding *SCN1A* PGVs in the MVP and UKB populations was unsurprising.

The low epilepsy penetrance of *SCN1A* PGVs was somewhat surprising, but since we detected <1.5% of all queried *SCN1A* PGVs across the 3 cohorts, the more deleterious PGVs were likely absent.

## Conclusion

Fortunately, most Veterans who carry PGVs in epilepsy genes will not develop epilepsy. Interestingly, we found that carriers of PGVs in AR-labeled genes had increased odds for epilepsy, suggesting inaccurate AR labels, latent compound heterozygosity, or that these PGVs may be risk alleles for adult-onset epilepsy. Our results underscore the importance of carefully evaluating genetic findings in the context of clinical, neurophysiological, age, and population-based phenotypes and not in isolation.

## Supporting information

STROBE checklist

***Supplementary Table ST1*** - “Ultrarare VAF<0.0001” aggregate analysis

***Supplementary Table ST2*** (supplement) – Rare (VAF<0.01) variant aggregate analysis

***Supplementary Table ST3*** - aggregate/summary data comparisons to UKB/GEL Stevelink data

***Supplementary Table ST4*** - MVP gene-level analyses of 170 genes

***Supplementary Table ST5 -*** Gene-level comparisons of MVP, UKB and GEL data for 91 overlapping genes

## Supplementary Materials

- Supplementary Methods (in Supplement word document)
- Supplementary Figures (2flow diagrams of gene/participant selection)
- Supplementary Tables (5 tables in Excel spreadsheet tabs and word document)
- MVP dbGAP manuscript accession number (phs001672)
- MVP Core Acknowledgements

## Acknowledgement list

- MVP Core Acknowledgement list

- The authors thank the MVP staff, researchers, and volunteers, who have contributed to MVP, and especially who previously served their country in the military and now generously agreed to enroll in the study (see mvp.va.gov for more information). The underlying work was based on data from the Million Veteran Program, Office of Research and Development, Veterans Heath Administration, and was supported by the Veterans Administration MVP award MVP000, MVP107 and MVP120. This work was supported using resources and facilities of the VHA Epilepsy Centers of Excellence (ECoE) and Department of Veterans Affairs (VA) Informatics and Computing Infrastructure (VINCI) which is funded under the research priority to Put VA Data to Work for Veterans (VA ORD 24-D4V-02), including data analytics by its Precision Medicine team and manuscript assistance by Kathryn Pridgen. Other contributions (not meeting criteria for authorship): MaryJo Pugh, Rizwana Rehman, Mayo O’Neal, Victoria Merritt, Nik Dembrow, Nishant Mishra, Colin Ellis, Alan Towne, and Ngoni Faya.

## MVP Funding and VA disclaimer statement

"This research is based on data from the MVP, Office of Research and Development, VHA, and was supported by MVP000, MVP107 and MVP120 as well as award #I21RX005453 (VHA Rehabilitation Research SPiRE) and Department of Defense (DoD) Congressionally Directed Medical Research Program (CDMRP) Epilepsy Research Program (ERP) PTERC award # EP240009. This publication does not represent the views of the Department of Veteran Affairs, Department of Defense, or the United States Government."

