## Supplementary material for "Pathogenic Epilepsy Gene Variant Prevalence and Penetrance Among U.S. Military Veterans in the Million Veteran Program Cohort": STROBE checklist

STROBE Statement—checklist of items that should be included in reports of observational studies

| | Item No. | Recommendation | Page No. | Relevant text from manuscript |
|---|---|---|---|---|
| **Title and abstract** | 1 | (*a*) Indicate the study's design with a commonly used term in the title or the abstract | 1 | "This retrospective cohort study…" |
| | | (*b*) Provide in the abstract an informative and balanced summary of what was done and what was found | 1-2 | See Abstract |
| **Introduction** | | | | |
| Background/rationale | 2 | Explain the scientific background and rationale for the investigation being reported | 3 | Studies have explored background polygenic risk, molecular consequence of the specific variants, and biobank population characteristics as probable contributors to the discrepancy, but factors such as mode of inheritance (MOI), single gene-level variation, and gene-disease relationship validity have not yet been explored. |
| Objectives | 3 | State specific objectives, including any prespecified hypotheses | 3 | Our primary aim was to determine the prevalence of PGVs in Veteran s and estimate the penetrance of epilepsy among carriers. We also evaluated 1) the effect of MOI and gene-disease relationship validity19,20 on prevalence and penetrance; 2) the relative prevalence and penetrance |

| | | | | |
|---|---|---|---|---|
|  |  |  |  | estimates in the MVP Veteran cohort compared to previous findings by Stevelink et al in the UKB/GEL cohorts3; and 3) single gene-level effects on prevalence and penetrance. |
| **Methods** | | | | |
| Study design | 4 | Present key elements of study design early in the paper | 4 | This study was performed within the MVP cohort and uses data from the VHA electronic health record (EHR)… Enrollment began in 2012 and is ongoing. EHR data are available for all participants who routinely received healthcare within the VHA system between 2002 and the administrative censoring date, September 30, 2024.” |
| Setting | 5 | Describe the setting, locations, and relevant dates, including periods of recruitment, exposure, follow-up, and data collection | 4 | “This study was performed withing the MVP cohort...” |
| Participants | 6 | (*a*) *Cohort study*—Give the eligibility criteria, and the sources and methods of selection of participants. Describe methods of follow-up<br>*Case-control study*—Give the eligibility criteria, and the sources and methods of case ascertainment and control selection. Give the rationale for the choice of cases and controls<br>*Cross-sectional study*—Give the eligibility criteria, and the sources and methods of selection of participants | 4 | “The primary analyses were performed within this WGS cohort.”<br>“An established VHA-validated algorithm identified epilepsy cases from VHA EHR data…” |
| | | (*b*) *Cohort study*—For matched studies, give matching criteria and number of exposed and unexposed | | |

| | | | | |
|---|---|---|---|---|
| | | *Case-control study*—For matched studies, give matching criteria and the number of controls per case | | |
| Variables | 7 | Clearly define all outcomes, exposures, predictors, potential confounders, and effect modifiers. Give diagnostic criteria, if applicable | 4, 5 | "The exposure was one or more epilepsy-associated rare variant, and outcome was an epilepsy diagnosis. Variables include sex, genetically inferred ancestry (GIA), and history of TBI or stroke." "MVP Phenotypes Definitions" section |
| Data sources/ measurement | 8* | For each variable of interest, give sources of data and details of methods of assessment (measurement). Describe comparability of assessment methods if there is more than one group | 4 | "TBI history and stroke history were determined by previously validated algorithms using EHR data." |
| Bias | 9 | Describe any efforts to address potential sources of bias | 5-6 | "We corrected for ascertainment bias in penetrance estimates using Bayes' theorem" |
| Study size | 10 | Explain how the study size was arrived at | n/a | Sample size determined by eligibility criteria |

Continued on next page

| | | | | |
|---|---|---|---|---|
| Quantitative variables | 11 | Explain how quantitative variables were handled in the analyses. If applicable, describe which groupings were chosen and why | Not applicable | |
| Statistical methods | 12 | (*a*) Describe all statistical methods, including those used to control for confounding | 5-6 | |
| | | (*b*) Describe any methods used to examine subgroups and interactions | 5-6 | Supplementary Figure 4 |
| | | (*c*) Explain how missing data were addressed | 11 | We presented all available data, but cannot account for variants/genes that were not found; this unquantified data missingness is a limitation of the study acknowledged in the discussion section. |
| | | (*d*) *Cohort study*—If applicable, explain how loss to follow-up was addressed<br>*Case-control study*—If applicable, explain how matching of cases and controls was addressed<br>*Cross-sectional study*—If applicable, describe analytical methods taking account of sampling strategy | 5-6, 11-12 | "limited in biobank studies due to immortal time bias" |
| | | (*e*) Describe any sensitivity analyses | n/a | n/a |
| **Results** | | | | |
| Participants | 13* | (a) Report numbers of individuals at each stage of study—eg numbers potentially eligible, examined for eligibility, confirmed eligible, included in the study, completing follow-up, and analysed | Supplement p. 3 | Supplementary Figure 1 |
| | | (b) Give reasons for non-participation at each stage | n/a | N/a |
| | | (c) Consider use of a flow diagram | Supplement p. 3 | Supplementary Figure 1 |
| Descriptive data | 14* | (a) Give characteristics of study participants (eg demographic, clinical, social) and information on exposures and potential confounders | 14 | Table 1. MVP participant characteristics |
| | | (b) Indicate number of participants with missing data for each variable of interest | 14 | Table 1. MVP participant characteristics |
| | | (c) *Cohort study*—Summarise follow-up time (eg, average and total amount) | 6, 14 | "Mean age at study enrollment was 65.6 years (standard deviation [SD] 13.0 yrs, range 20-101), mean age at censorship or death was 74.6 (SD 12.3, range 26-108)" |

| | | | | |
|---|---|---|---|---|
| Outcome data | 15* | *Cohort study*—Report numbers of outcome events or summary measures over time | 6-7 | "MVP Epilepsy Prevalence and Penetrance" |
| | | *Case-control study*—Report numbers in each exposure category, or summary measures of exposure | | |
| | | *Cross-sectional study*—Report numbers of outcome events or summary measures | | |
| Main results | 16 | (*a*) Give unadjusted estimates and, if applicable, confounder-adjusted estimates and their precision (eg, 95% confidence interval). Make clear which confounders were adjusted for and why they were included | 6-8 | "Refuted, disputed and/or limited genes were not associated with epilepsy"<br>"Penetrance of AD-labeled ultrarare PGVs ranged between 7.5–12.8% depending on gene grouping"<br>"Carriers of autosomal recessive ultrarare PGVs had increased odds of epilepsy diagnosis" |
| | | (*b*) Report category boundaries when continuous variables were categorized | n/a | |
| | | (*c*) If relevant, consider translating estimates of relative risk into absolute risk for a meaningful time period | n/a | |

Continued on next page

| | | | | |
|---|---|---|---|---|
| Other analyses | 17 | Report other analyses done—eg analyses of subgroups and interactions, and sensitivity analyses | 8 | "Comparison of MVP, UKB and GEL Cohort Prevalence and Penetrance" |
| **Discussion** | | | | |
| Key results | 18 | Summarise key results with reference to study objectives | 9 | "Approximately 1% of Veterans with WGS in MVP carried ultrarare PGVs in epilepsy-associated genes…" |
| Limitations | 19 | Discuss limitations of the study, taking into account sources of potential bias or imprecision. Discuss both direction and magnitude of any potential bias | 11-13 | Limitations discussed throughout the Discussion |
| Interpretation | 20 | Give a cautious overall interpretation of results considering objectives, limitations, multiplicity of analyses, results from similar studies, and other relevant evidence | 9-11 | See Discussion |
| Generalisability | 21 | Discuss the generalisability (external validity) of the study results | 11 | "Penetrance estimates were low for AD-labeled PGVs in the MVP cohort, but fell within range of those in the UKB and GEL cohorts and other similar population-based studies, thus validating Stevelink et al's findings…" |
| **Other information** | | | | |
| Funding | 22 | Give the source of funding and the role of the funders for the present study and, if applicable, for the original study on which the present article is based | 22 | "This research is based on data from the MVP, Office of Research and Development, VHA, and was supported by MVP000, MVP107 and MVP120 as well as award #I21RX005453 (VHA Rehabilitation Research SPiRE) and Department of Defense (DoD) Congressionally Directed Medical Research Program (CDMRP) |

Epilepsy Research Program (ERP)
PTERC award # EP240009.”

*Give information separately for cases and controls in case-control studies and, if applicable, for exposed and unexposed groups in cohort and cross-sectional studies.

**Note:** An Explanation and Elaboration article discusses each checklist item and gives methodological background and published examples of transparent reporting. The STROBE checklist is best used in conjunction with this article (freely available on the Web sites of PLoS Medicine at http://www.plosmedicine.org/, Annals of Internal Medicine at http://www.annals.org/, and Epidemiology at http://www.epidem.com/). Information on the STROBE Initiative is available at www.strobe-statement.org.
