## Supplement for "Pathogenic Epilepsy Gene Variant Prevalence and Penetrance Among U.S. Military Veterans in the Million Veteran Program Cohort"

<u>Supplement outline</u>:

S1: MVP WGS Pipeline details

S2: Flow diagrams of case and gene selection

S3: UKB/GEL study detail summary

S4: Penetrance estimate formula

S5: Lay summary of study

S6: VA MVP Core Acknowledgements for Publications (October 2025)

Supplement-specific citations

<u>Supplementary tables</u>:

***Supplementary Table ST1:*** "Ultrarare VAF<0.0001" aggregate analysis

***Supplementary Table ST2*** : Rare (VAF<0.01) variant aggregate analysis

***Supplementary Table ST3:*** aggregate/summary data comparisons to UKB/GEL Stevelink data

***Supplementary Table ST4:*** MVP gene-level analyses of 170 genes

***Supplementary Table ST5:*** Gene-level comparisons of MVP, UKB and GEL data for 91 overlapping genes

### <u>S1: MVP WGS pipeline details</u>

To identify DNA variations within the sequenced human genomes, a modified version of the Broad Institute's $5 GATK workflow (available at https://github.com/gatk-workflows/five-dollar-genome-analysis-pipeline) was used[1]. The GRCh38 human reference genome served as the foundation for alignment and variant identification. BWA-MEM (version 0.7.15) conducted the sequence alignment process, with results subsequently converted to CRAM format through lossless compression[2]. The identification of variants employed GATK version 4.1.0.0 utilizing the haplotypeCaller tool, which produced gVCF files encompassing reference call blocks, single nucleotide variants (SNVs)[3]. Quality control was implemented across several stages. For sequencing assessment, FastQC (version 0.11.4) evaluated base sequence quality metrics, read quality parameters, and GC content; for alignment evaluation, SAMTools flagstat (version 0.1.19) determined the count and proportion of correctly mapped and paired reads[4]. Regarding variant calling outputs, RTG Tools vcfstats (version 3.7.1) quantified SNVs per genome, calculated the transition-to-transversion ratio for SNVs, and determined the ratio between heterozygous and homozygous SNV genotypes[5]. Furthermore, verifybamID within GATK 4.1.0.0 was employed to assess DNA contamination levels for individual genomes[3].

S2: UKB/GEL study detail summary[6]

Mean age at enrollment was reported as 30 years (range 0-99) in the GEL cohort and enrollment age range was between 40-69 years in the UKB cohort. Sex and ancestry for the UK-based cohorts were not reported. In both UK-based cohorts, germline WGS was performed using Illumina next generation sequencing with a mean depth of 32x. Variants were called using a DRAGEN platform in UKB and iSAAC in GEL[7]. In GEL, a large proportion of epilepsy cases were recruited for “early onset or syndromic epilepsy”. In the UKB, the authors used “an age at onset cutoff <18 years of age to exclude acquired causes of epilepsy”. Phenotype definition: In UKB and GEL, epilepsy is defined using epilepsy-related diagnostic ICD codes[6]; no concurrent medication prescriptions were required/available in contrast to the MVP epilepsy phenotype definition. Genotype definition: the authors queried “all pathogenic variants in AD epilepsy genes in ClinVar [in the UKB and GEL cohorts], yielding 4294 ultrarare variants across 92 DEE genes”[6]. The authors report performing manual curation of certain variants to ensure association of the specific variant with DEE. In UKB, they detected 111 unique PGVs across 33 genes in 294 carriers. In GEL, they detected 37 unique variants across 15 genes in 45 carriers.

S3: Penetrance estimate formula

$$P(D|G) = \frac{P(G|D) \times P(D)}{P(G|D) \times P(D) + P(G|\overline{D}) \times (1 - P(D))}$$

where D = disease, G = genotype (carrying a pathogenic variant), and $\overline{D})$ = absence of disease. P(D|G) is the penetrance (probability of disease given a genotype); P(G|D) is the genotype frequency in cases; $P(G|\overline{D})$ is the genotype frequency in controls, and P(D) is the general population prevalence of epilepsy[6].

S4 Flow diagrams of case and gene selection:

A) Cohort selection:

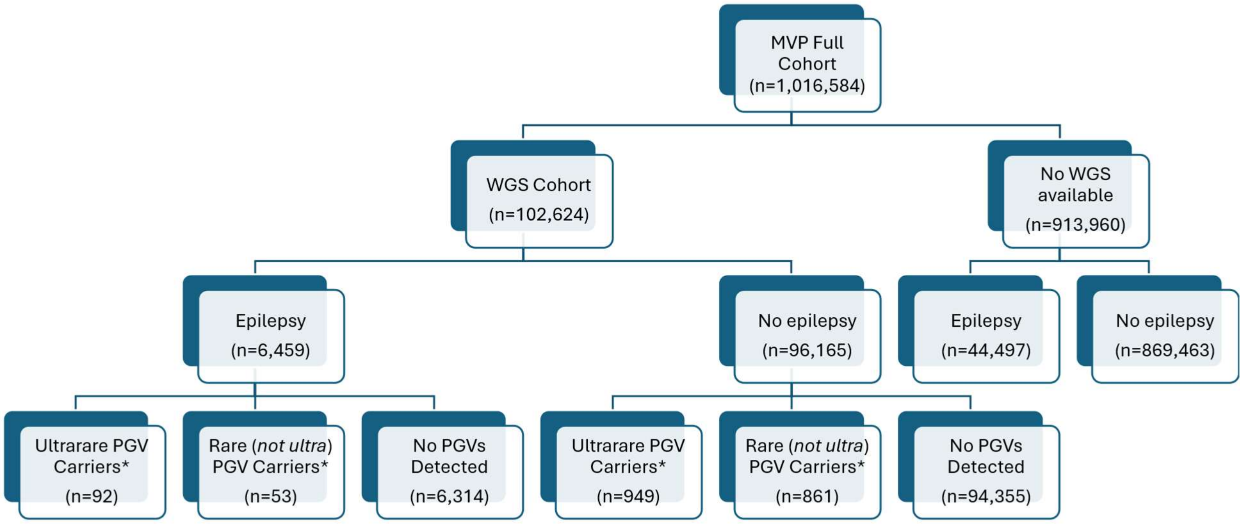


*= Excludes limited, refuted, disputed genes

B) Gene selection by ClinGen classification:

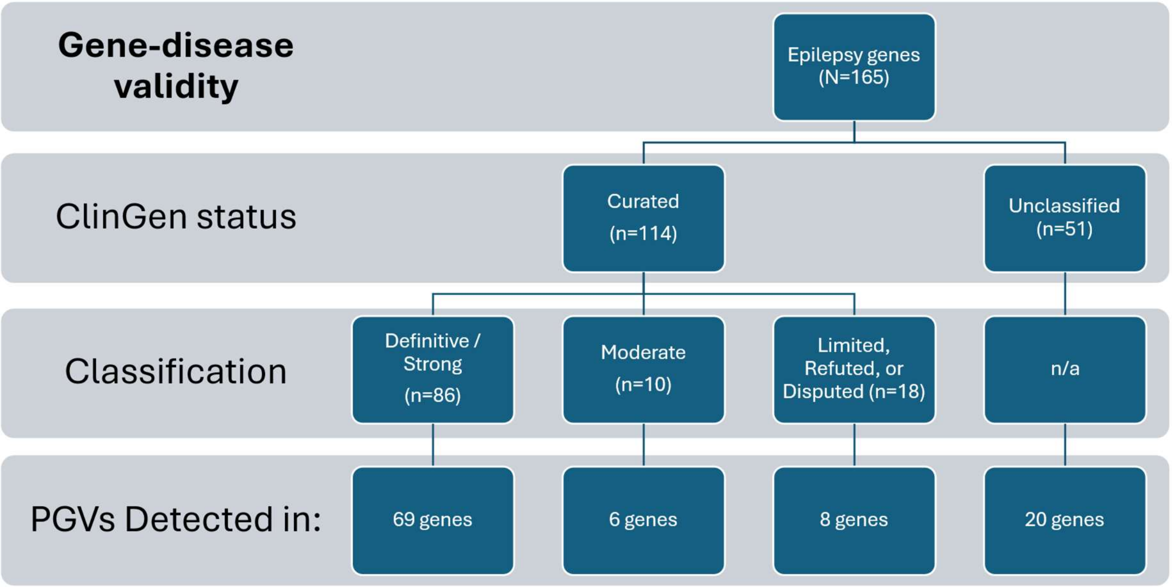

C) Gene selection by MOI:

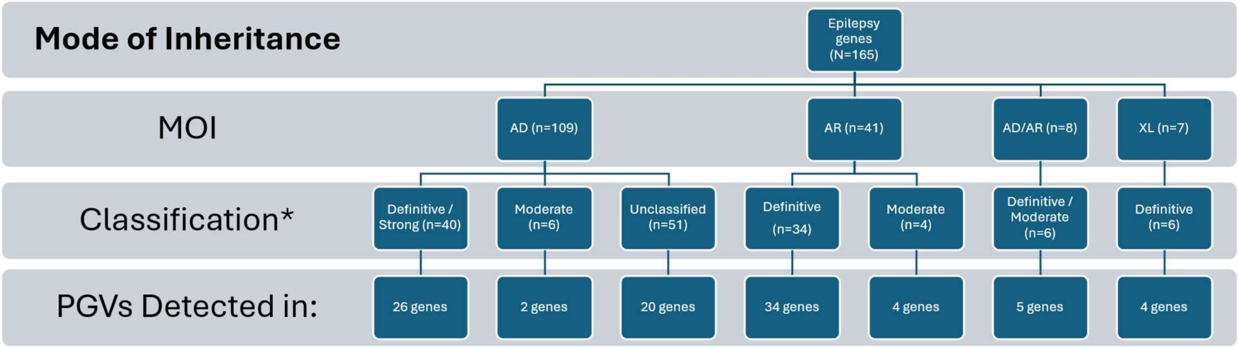


*= Excludes limited, refuted, disputed genes – which were not used in MOI analyses

<u>S5: Lay summary of study</u>

Background and Objectives: Genetic causes of epilepsy are well-established in children but not well understood in adults. There are few studies of epilepsy genetics in older adults, U.S. Veterans, and people with acquired causes of epilepsy like traumatic brain injury (TBI) and stroke. To address this, we determined the prevalence of pathogenic germline variants (PGVs, i.e. rare genetic changes in our DNA) in known epilepsy-associated genes in an ancestrally diverse cohort of older Veterans and examined the prevalence of epilepsy among carriers compared to other cohorts.

Methods: This retrospective cohort study used electronic health record (EHR) data from Veterans enrolled in the Million Veteran Program (MVP) biobank who had whole genome sequencing (WGS) data available. We identified Veterans with ≥1 pathogenic/likely pathogenic single nucleotide variants (SNVs) within one or more of 165 expert-curated epilepsy genes.

Results: There were 102,624 MVP participants with WGS data. Mean age at censorship or death was 74.6 years, and 6.1% were female. Among the cohort, 1,955 participants (1.91%) carried ≥1 rare PGVs and 1,041 participants (1.01%) carried ultrarare PGVs. Most carriers were not diagnosed with epilepsy, though ultrarare PGV carriers had 1.45-1.83 increased odds of an epilepsy diagnosis compared to non-carriers.

Discussion: Around 90% of Veterans carrying ultrarare PGVs in epilepsy genes were not diagnosed with epilepsy, although they were at increased risk for epilepsy compared to non-carriers.

S6: VA Million Veteran Program Core Acknowledgements for Publications (October 2025)

MVP Program Office

- Sumitra Muralidhar, Ph.D., Program Director

US Department of Veterans Affairs, 810 Vermont Avenue NW, Washington, DC 20420

- Jennifer Moser, Ph.D., Associate Director, Scientific Programs

US Department of Veterans Affairs, 810 Vermont Avenue NW, Washington, DC 20420

- Jennifer E. Deen, B.S., Associate Director, Cohort & Public Relations

US Department of Veterans Affairs, 810 Vermont Avenue NW, Washington, DC 20420

MVP Steering Committee

- Co-Chair: Philip S. Tsao, Ph.D.

VA Palo Alto Health Care System, 3801 Miranda Avenue, Palo Alto, CA 94304

- Co-Chair: Sumitra Muralidhar, Ph.D.

US Department of Veterans Affairs, 810 Vermont Avenue NW, Washington, DC 20420

- J. Michael Gaziano, M.D., M.P.H.

VA Boston Healthcare System, 150 S. Huntington Avenue, Boston, MA 02130

- Adriana Hung, M.D., M.P.H.,

VA Tennessee Valley Healthcare System, 1310 24th Avenue, South Nashville, TN 37212

- Dave Oslin, M.D.

Philadelphia VA Medical Center, 3900 Woodland Avenue, Philadelphia, PA 19104

- Deepak Voora, M.D.

Durham VA Medical Center, 508 Fulton Street, Durham, NC 27705

MVP Co-Principal Investigators

- J. Michael Gaziano, M.D., M.P.H.

VA Boston Healthcare System, 150 S. Huntington Avenue, Boston, MA 02130

- Philip S. Tsao, Ph.D.

VA Palo Alto Health Care System, 3801 Miranda Avenue, Palo Alto, CA 94304

MVP Core Operations

- Jessica V. Brewer, M.P.H., Director, MVP Cohort Operations

VA Boston Healthcare System, 150 S. Huntington Avenue, Boston, MA 02130

- Mary T. Brophy M.D., M.P.H., Director, VA Central Biorepository

VA Boston Healthcare System, 150 S. Huntington Avenue, Boston, MA 02130

- Kelly Cho, M.P.H, Ph.D., Director, MVP Phenomics

VA Boston Healthcare System, 150 S. Huntington Avenue, Boston, MA 02130

- Lori Churby, B.S., Director, MVP Regulatory Affairs

VA Palo Alto Health Care System, 3801 Miranda Avenue, Palo Alto, CA 94304

- Jacob T. Kean, Ph.D., Acting Director, VA Informatics and Computing Infrastructure (VINCI)

MVP Core Acknowledgements for Publications_October 2025

VA Salt Lake City Health Care System, 500 Foothill Drive, Salt Lake City, UT 84148

- Saiju Pyarajan Ph.D., Director, Data and Computational Sciences

VA Boston Healthcare System, 150 S. Huntington Avenue, Boston, MA 02130

- Robert Ringer, Pharm.D., Director, VA Albuquerque Central Biorepository

New Mexico VA Health Care System, 1501 San Pedro Drive SE, Albuquerque, NM 87108

- Luis E. Selva, Ph.D., Director, MVP Biorepository Coordination

VA Boston Healthcare System, 150 S. Huntington Avenue, Boston, MA 02130

- Shahpoor (Alex) Shayan, M.S., Director, MVP PRE Informatics

VA Boston Healthcare System, 150 S. Huntington Avenue, Boston, MA 02130

- Brady Stephens, M.S., Principal Investigator, MVP Information Center

Canandaigua VA Medical Center, 400 Fort Hill Avenue, Canandaigua, NY 14424

- Stacey B. Whitbourne, Ph.D., Director, MVP Cohort Development and Management

VA Boston Healthcare System, 150 S. Huntington Avenue, Boston, MA 0213

Supplement-specific citations

1. Ross, P. B., Song, J., Tsao, P. S. & Pan, C. Trellis for efficient data and task management in the VA Million Veteran Program. *Sci. Rep.* **11**, 23229 (2021).
2. Regier, A. A. *et al.* Functional equivalence of genome sequencing analysis pipelines enables harmonized variant calling across human genetics projects. *Nat. Commun.* **9**, 4038 (2018).
3. McKenna, A. *et al.* The Genome Analysis Toolkit: a MapReduce framework for analyzing next-generation DNA sequencing data. *Genome Res.* **20**, 1297–1303 (2010).
4. Danecek, P. *et al.* Twelve years of SAMtools and BCFtools. *GigaScience* **10**, giab008 (2021).
5. Danecek, P. *et al.* The variant call format and VCFtools. *Bioinformatics* **27**, 2156–2158 (2011).
6. Stevelink, R. *et al.* Penetrance of pathogenic epilepsy variants is low and shaped by common genetic background. *Epilepsia* https://doi.org/10.1111/epi.70018 (2025) doi:10.1111/epi.70018.
7. AggV2 sample QC - Genomics England Research Environment User Guide. https://re-docs.genomicsengland.co.uk/sample_qc/.

Supplementary Tables:

***Supplementary Table ST1:*** "Ultrarare VAF<0.0001" aggregate analysis

| | All genes | All: except limited/ disputed / refuted | All: definitive | All: limited / disputed / refuted | AD: all | AD: except limited/ disputed / refuted | MALE only- AD: except lim/dis/ref | AD: Not classified (Stevelink) | AD: definitive | AD: Moderate | AD: limited, disputed or refuted | AR: all | AR: definitive | AR: Moderate | AR: except limited | MALE only - AR: except limited | AD/AR: all | AD/AR: except disputed /refuted | XL: all except disputed/ refuted |
|---|---|---|---|---|---|---|---|---|---|---|---|---|---|---|---|---|---|---|---|
| Genes attempted | 165 | 147 | 86 | 18 | 109 | 97 | 97 | 51 | 40 | 6 | 12 | 41 | 34 | 4 | 38 | 38 | 8 | 6 | 7 |
| Variants attempted | 9847 | 9468 | 7468 | 379 | 7526 | 7181 | 7181 | 1815 | 5264 | 102 | 345 | 1665 | 1506 | 83 | 1636 | 1636 | 263 | 260 | 440 |
| Genes detected in all MVP | 102 | 94 | 68 | 8 | 51 | 47 | 44 | 20 | 25 | 2 | 4 | 40 | 34 | 4 | 38 | 38 | 7 | 5 | 4 |
| Variants detected in MVP | 570 | **517** | 446 | 53 | 179 | 132 | 122 | 38 | 75 | 19 | 47 | 360 | 343 | 14 | 357 | 349 | 15 | 12 | 16 |
| % variants detected/attempted MVP | 0.0579 | 0.0546 | 0.0597 | 0.1398 | 0.0238 | 0.0184 | 0.0170 | 0.0209 | 0.0142 | 0.1863 | 0.1362 | 0.2162 | 0.2278 | 0.1687 | 0.2182 | 0.2133 | 0.0570 | 0.0462 | 0.0364 |
| Carriers (all) in MVP | 1149 | **1041** | 904 | **110** | 298 | **203** | 187 | 57 | 96 | 50 | 95 | 807 | 773 | 31 | **804** | 751 | 31 | **19** | **20** |
| Affected carriers (epilepsy) in MVP | 95 | 92 | 75 | 3 | 24 | 21 | 19 | 9 | 9 | 3 | 3 | 71 | 65 | 6 | 71 | 65 | 1 | 1 | 0 |
| Proportion w epi MVP (affected/carriers: aka prevalence of epilepsy among carriers) | 0.0827 | **0.0884** | 0.0830 | 0.0273 | 0.0805 | **0.1034** | 0.1016 | 0.1579 | 0.0938 | 0.0600 | 0.0316 | 0.0880 | 0.0841 | 0.1935 | **0.0883** | 0.0866 | 0.0323 | 0.0526 | **0.0000** |
| Variant frequency in all MVP | 0.0112 | **0.0101** | 0.0088 | 0.0011 | 0.0029 | **0.0020** | 0.0018 | 0.0006 | 0.0009 | 0.0005 | 0.0009 | 0.0079 | 0.0075 | 0.0003 | **0.0078** | 0.0073 | 0.0003 | 0.0002 | **0.0002** |
| Variant frequency in epilepsy in MVP | 0.0147 | 0.0142 | 0.0116 | 0.0005 | 0.0037 | 0.0033 | 0.0029 | 0.0014 | 0.0014 | 0.0005 | 0.0005 | 0.0110 | 0.0101 | 0.0009 | 0.0110 | 0.0101 | 0.0002 | 0.0002 | 0.0000 |
| Variant frequency in *no* epilepsy in MVP | 0.0110 | 0.0099 | 0.0086 | 0.0011 | 0.0028 | 0.0019 | 0.0017 | 0.0005 | 0.0009 | 0.0005 | 0.0010 | 0.0077 | 0.0074 | 0.0003 | 0.0076 | 0.0071 | 0.0003 | 0.0002 | 0.0002 |
| MVP PENETRANCE: after applying correction formula (using 0.7% as epilepsy prevalence) | 0.0094 | 0.0101 | 0.0094 | 0.0029 | 0.0091 | 0.0120 | 0.0117 | 0.0193 | 0.0107 | 0.0067 | 0.0034 | 0.0100 | 0.0095 | 0.0246 | 0.0100 | 0.0098 | 0.0035 | 0.0058 | 0.0000 |
| MVP PENETRANCE: after applying correction formula (using 5% as epilepsy prevalence) | 0.0660 | **0.0706** | 0.0662 | 0.0215 | 0.0642 | **0.0829** | 0.0814 | 0.1281 | **0.0750** | 0.0476 | 0.0249 | 0.0703 | 0.0671 | 0.1583 | **0.0705** | 0.0691 | 0.0254 | 0.0417 | **0.0000** |
| Odds of epilepsy in carriers vs non-carriers (OR) | 1.35 | **1.45** | 1.35 | **0.42** | 1.31 | **1.72** | 1.69 | **2.79** | **1.54** | 0.95 | 0.49 | 1.44 | 1.37 | 3.58 | **1.45** | 1.41 | 0.5 | 0.83 | n/a (no cas |
| 95% CI | *1.09-1.66* | *1.17-1.80* | *1.07-1.71* | 0.13-1.31 | 0.286-1.9 | 1.09-2.70 | 1.04-2.71 | 1.37-16.64 | 0.76 – 3.06 | 0.30-3.05 | 0.15-1.53 | 1.13 – 1.84 | 1.06 – 1.77 | 1.47-8.72 | 1.13-1.85 | 1.10-1.83 | 0.07-3.64 | 0.11-6.20 | n/a (no cas |
| Female carriers (%) | 0.0661 | **0.0663** | 0.0653 | 0.0636 | 0.0772 | **0.0788** | n/a | 0.0702 | 0.0833 | 0.0800 | 0.0737 | 0.0657 | 0.0660 | 0.0645 | **0.0659** | n/a | 0.0000 | 0.0000 | **0.0000** |
| Carriers mean age (SD) (at enrollment) | 65.1 (13.0) | 65.0 (12.9) | 65.1 (12.9) | 65.2 (12.9) | 64.4 (13.4 | 63.9 (13.2 | 65.3 (12.4 | 63.0 (15.3) | 63.7 (12.7) | 65.4 (11.3 | 65.3 (13.8) | 65.2 (12.8 | 65.3 (12.9 | 64.9 (9.8) | 65.2 (12.8 | 66.0 (12.5 | 65.0 (13.6 | 64.4 (14.2 | 66.6 (13.8) |
| Carriers median age (range) (at enrollment) | 65.0 (24.0-99 | 65.0 (24.0-9 | 65.0 (24.0-9 | 65.0 (29.0-9 | 64.0 (27.0 | 64.0 (27.0 | 64.0 (32.0 | 63.0 (29.0-9 | 64.0 (27.0- | 64.0 (43.0- | 65.0 (29.0- | 65.0 (24.0- | 65.0 (24.0- | 64.0 (43.0- | 65.0 (24.0- | 66.0 (24.0- | 65.0 (31.0- | 65.0 (31.0- | 68.5 (33.0- |
| **Age <40 yrs (%) (at enrollment)** | 0.0374 | 0.0355 | 0.0354 | 0.0545 | 0.0470 | 0.0443 | 0.0267 | 0.0877 | 0.0417 | 0.0000 | 0.0526 | 0.0335 | 0.0336 | 0.0000 | 0.0323 | 0.0280 | 0.0323 | 0.0526 | 0.0500 |
| 40-<65 yrs (%) (at enrollment) | 0.4404 | 0.4438 | 0.4325 | 0.4182 | 0.4765 | 0.5074 | 0.4973 | 0.4737 | 0.4896 | 0.5800 | 0.4105 | 0.4312 | 0.4282 | 0.5161 | 0.4316 | 0.4115 | 0.4516 | 0.4211 | 0.3500 |
| 65+yrs (%) (at enrollment) | 0.5222 | 0.5207 | 0.5321 | 0.5273 | 0.4765 | **0.4483** | 0.4759 | 0.4386 | 0.4688 | 0.4200 | 0.5368 | 0.5353 | 0.5382 | 0.4839 | **0.5361** | 0.5606 | 0.5161 | 0.5263 | 0.6000 |
| Carriers mean age (SD) (at censorship) | 74.2 (12.3) | 74.2 (12.2) | 74.3 (12.3) | 74.2 (13.6) | 73.5 (12.6 | 73.2 (12.4 | 74.5 (11.5 | 72.2 (14.4) | 73.0 (12.2) | 74.6 (10.4 | 74.2 (13.1) | 74.4 (12.1 | 74.4 (12.2 | 74.1 (8.9) | 74.4 (12.1 | 75.1 (11.8 | 74.6 (13.9 | 74.2 (13.9 | 76.9 (14.2) |
| Carriers median age (range) (at censorship) | 75.0 (31.4-10 | 75.0 (31.4-1 | 75.0 (31.4-1 | 76.1 (37.1-1 | 74.4 (34.0 | 73.3 (34.0 | 74.4 (44.2 | 71.2 (38.1, 9 | 74.0 (34.0- | 73.3 (54.8- | 76.0 (37.1- | 75.0 (31.4- | 75.0 (31.4- | 73.3 (53.1- | 75.0 (31.4- | 75.4 (31.4- | 75.3 (40.6- | 75.2 (40.6- | 79.1 (43.6- |
| **Age <40 yrs (%) (at censorship)** | 0.0078 | 0.0058 | 0.0055 | 0.0273 | 0.0134 | 0.0099 | 0.0000 | 0.0175 | 0.0104 | 0.0000 | 0.0211 | 0.0062 | 0.0052 | 0.0000 | 0.0050 | 0.0053 | 0.0000 | 0.0000 | 0.0000 |
| 40-<65 yrs (%) (at censorship) | 0.1862 | 0.1883 | 0.1914 | 0.1818 | 0.1946 | 0.2020 | 0.1658 | 0.2456 | 0.2083 | 0.1400 | 0.1789 | 0.1846 | 0.1889 | 0.0968 | 0.1853 | 0.1598 | 0.2258 | 0.2105 | 0.2000 |
| 65+yrs (%) (at censorship) | 0.8059 | 0.8060 | 0.8031 | 0.7909 | 0.7919 | 0.7882 | 0.8342 | 0.7368 | 0.7813 | 0.8600 | 0.8000 | 0.8092 | 0.8060 | 0.9032 | 0.8097 | 0.8349 | 0.7742 | 0.7895 | 0.8000 |
| Carrier Ancestry | | | | | | | | | | | | | | | | | | | |
| **EUR (%)** | 0.6867 | 0.6878 | 0.6803 | 0.6636 | 0.6846 | **0.6995** | 0.7112 | 0.6491 | 0.6875 | 0.7800 | 0.6526 | 0.6877 | 0.6805 | 0.8387 | **0.6866** | 0.6951 | 0.6129 | 0.5789 | **0.7500** |
| **AFR (%)** | 0.2341 | 0.2334 | 0.2356 | 0.2545 | 0.2483 | **0.2463** | 0.2513 | 0.2982 | 0.2292 | 0.2200 | 0.2526 | 0.2305 | 0.2380 | 0.0645 | **0.2313** | 0.2210 | 0.2581 | 0.2105 | **0.2000** |
| AMR (%) | 0.0618 | 0.0624 | 0.0653 | 0.0545 | 0.0503 | 0.0443 | 0.0267 | 0.0526 | 0.0625 | 0.0000 | 0.0632 | 0.0632 | 0.0621 | 0.0968 | 0.0634 | 0.0652 | 0.1290 | 0.2105 | 0.0500 |
| EAS | 0.0070 | 0.0067 | 0.0077 | 0.0091 | 0.0067 | 0.0049 | 0.0053 | 0.0000 | 0.0104 | 0.0000 | 0.0105 | 0.0074 | 0.0078 | 0.0000 | 0.0075 | 0.0067 | 0.0000 | 0.0000 | 0.0000 |
| SAS | 0.0009 | 0.0010 | 0.0011 | 0.0000 | 0.0000 | 0.0000 | 0.0000 | 0.0000 | 0.0000 | 0.0000 | 0.0000 | 0.0012 | 0.0013 | 0.0000 | 0.0012 | 0.0013 | 0.0000 | 0.0000 | 0.0000 |
| Other | 0.0096 | 0.0086 | 0.0100 | 0.0182 | 0.0101 | 0.0049 | 0.0053 | 0.0000 | 0.0104 | 0.0000 | 0.0211 | 0.0099 | 0.0103 | 0.0000 | 0.0100 | 0.0107 | 0.0000 | 0.0000 | 0.0000 |

### ***Supplementary Table ST2:*** Rare (VAF<0.01) variant aggregate analysis

| | All genes | All: except limited/ disputed / refuted | All: definitive | All: limited / disputed / refuted | AD: all | AD: except limited/ disputed / refuted | MALE only- AD: except lim/dis/ref | AD: Not classified (Stevelink) | AD: definitive | AD: Moderate | AD: limited, disputed or refuted | AR: all | AR: definitive | AR: Moderate | AR: except limited | MALE only - AR: except limited | AD/AR: all | AD/AR: except disputed /refuted | XL: all |
|---|---|---|---|---|---|---|---|---|---|---|---|---|---|---|---|---|---|---|---|
| Genes attempted | 165 | 147 | 86 | 18 | 109 | 97 | 97 | 51 | 40 | 6 | 12 | 41 | 34 | 4 | 38 | 38 | 8 | 6 | 7 |
| Variants attempted | 9847 | 9468 | 7468 | 379 | 7526 | 7181 | 7181 | 1815 | 5264 | 102 | 345 | 1665 | 1506 | 83 | 1636 | 1636 | 263 | 260 | 440 |
| Genes detected in all MVP | 103 | 95 | 69 | 8 | 52 | 48 | 45 | 20 | 26 | 2 | 4 | 40 | 34 | 4 | 38 | 38 | 7 | 5 | 4 |
| Variants detected in MVP | **594** | **541** | 468 | 53 | 180 | 133 | 123 | 38 | 76 | 19 | 47 | 382 | 363 | 16 | 379 | 371 | 16 | 13 | 16 |
| % variants detected/attempted MVP | 0.0603 | 0.0571 | 0.0627 | 0.1398 | 0.0239 | 0.0185 | 0.0171 | 0.0209 | 0.0144 | 0.1863 | 0.1362 | 0.2294 | 0.2410 | 0.1928 | 0.2317 | 0.2268 | 0.0608 | 0.0500 | 0.0364 |
| **Carriers (all) in MVP** | **2063** | **1955** | **1638** | **110** | **317** | **222** | **206** | **57** | **115** | **50** | **95** | **1692** | **1478** | **212** | **1689** | **1591** | **43** | **31** | **20** |
| Affected carriers (epilepsy) in MVP | 148 | 145 | 116 | 3 | 24 | 21 | 19 | 9 | 9 | 3 | 3 | 124 | 106 | 18 | 124 | 117 | 1 | 1 | 0 |
| **Proportion w epi MVP (affected/carriers: aka prevalence of epilepsy among carriers)** | **0.0717** | **0.0742** | **0.0708** | **0.0273** | **0.0757** | **0.0946** | **0.0922** | **0.1579** | **0.0783** | **0.0600** | **0.0316** | **0.0733** | **0.0717** | **0.0849** | **0.0734** | **0.0735** | **0.0233** | **0.0323** | **0.0000** |
| Variant frequency in all MVP | 0.0201 | **0.0191** | 0.0160 | 0.0011 | 0.0031 | 0.0022 | 0.0020 | 0.0006 | 0.0011 | 0.0005 | 0.0009 | 0.0165 | 0.0144 | 0.0021 | 0.0165 | 0.0155 | 0.0004 | 0.0003 | 0.0002 |
| Variant frequency in epilepsy in MVP | 0.0229 | **0.0224** | 0.0180 | 0.0005 | 0.0037 | 0.0033 | 0.0029 | 0.0014 | 0.0014 | 0.0005 | 0.0005 | 0.0192 | 0.0164 | 0.0028 | 0.0192 | 0.0181 | 0.0002 | 0.0002 | 0.0000 |
| Variant frequency in *no* epilepsy in MVP | 0.0199 | 0.0188 | 0.0158 | 0.0011 | 0.0030 | 0.0021 | 0.0019 | 0.0005 | 0.0011 | 0.0005 | 0.0010 | 0.0163 | 0.0143 | 0.0020 | 0.0163 | 0.0153 | 0.0004 | 0.0003 | 0.0002 |
| MVP PENETRANCE: after applying correction formula (using 0.7% as epilepsy prevalence) | 0.0080 | 0.0083 | 0.0079 | 0.0029 | 0.0085 | 0.0108 | 0.0106 | 0.0193 | 0.0088 | 0.0067 | 0.0034 | 0.0082 | 0.0080 | 0.0096 | 0.0082 | 0.0083 | 0.0025 | 0.0035 | 0.0000 |
| **MVP PENETRANCE: after applying correction formula (using 5% as epilepsy prevalence)** | **0.0571** | **0.0591** | **0.0564** | **0.0215** | **0.0603** | **0.0757** | **0.0737** | **0.1281** | **0.0624** | **0.0476** | **0.0249** | **0.0584** | **0.0571** | **0.0678** | **0.0585** | **0.0586** | **0.0183** | **0.0255** | **0.0000** |
| **Odds of epilepsy in carriers vs non-carriers (OR)** | **1.15** | **1.12** | **1.14** | **0.42** | **1.22** | **1.56** | **1.51** | **2.79** | **1.26** | **0.95** | **0.49** | **1.18** | **1.15** | **1.38** | **1.18** | **1.19** | **0.35** | **0.50** | **n/a (no cases)** |
| 95% CI | 0.97-1.37 | 0.94-1.32 | 0.94-1.38 | 0.13-1.31 | 0.80-1.85 | 0.99-2.44 | 0.94-2.43 | **1.37-16.64** | 0.64-2.50 | 0.30-3.05 | 0.15-1.53 | 0.98-1.42 | 0.94-1.41 | 0.85-2.24 | 0.98-1.42 | 0.98-1.43 | 0.05-2.58 | 0.07-3.64 | n/a (no cases) |
| Female carriers (%) | 0.0591 | 0.0588 | 0.0580 | 0.0636 | 0.0726 | 0.0721 | n/a | 0.0702 | 0.0696 | 0.0800 | 0.0737 | 0.0579 | 0.0582 | 0.0566 | 0.0580 | n/a | 0.0233 | 0.0323 | 0.0000 |
| Carriers mean age (SD) (at enrollment) | 65.7 (12.8) | 65.7 (12.7 | 65.7 (12.7 | 65.2 (12.7) | 64.6 (13.6 | 64.3 (13.5 | 65.5 (12.8 | 63.0 (15.3) | 64.4 (13.4 | 65.4 (11.3 | 65.3 (13.8 | 65.9 (12.6 | 65.8 (12.6 | 66.5 (12.7 | 65.9 (12.6 | 66.6 (12.2 | 65.1 (13.3 | 64.7 (13.6 | 66.6 (13.8) |
| Carriers median age (range) (at enrollment) | 65.0 (24.0-99.0 | 65.0(24.0- | 65.0 (24.0- | 65.0 (29.0-0 | 64.0 (24.0- | 64.0 (24.0- | 64.0 (24.0- | 63.0 (29.0-9 | 64.0 (24.0- | 64.0 (43.0- | 65.0 (29.0- | 65.0 (24.0- | 65.0 (24.0- | 66.0 (30.0- | 65.0 (24.0- | 66.0 (24.0- | 64.0 (31.0- | 64.0 (31.0- | 68.5 (33.0-86.0 |
| **Age <40 yrs (%) (at enrollment)** | 0.0344 | 0.0332 | 0.0317 | 0.0545 | 0.0473 | 0.0450 | 0.0291 | 0.0877 | 0.0435 | 0.0000 | 0.0526 | 0.0319 | 0.0304 | 0.0377 | 0.0314 | 0.0245 | 0.0233 | 0.0323 | 0.0500 |
| 40-<65 yrs (%) (at enrollment) | 0.4232 | 0.4240 | 0.4206 | 0.4182 | 0.4732 | 0.5000 | 0.4903 | 0.4737 | 0.4783 | 0.5800 | 0.4105 | 0.4137 | 0.4154 | 0.4009 | 0.4139 | 0.3979 | 0.5116 | 0.5161 | 0.3500 |
| 65+yrs (%) (at enrollment) | 0.5424 | 0.5427 | 0.5476 | 0.5273 | 0.4795 | 0.4550 | 0.4806 | 0.4386 | 0.4783 | 0.4200 | 0.5368 | 0.5550 | 0.5541 | 0.5613 | 0.5548 | 0.5776 | 0.4651 | 0.4516 | 0.6000 |
| Carriers mean age (SD) (at censorship) | 74.9 (12.1) | 75.0 (12.0 | 75.0 (12.0 | 74.2 (13.6) | 73.7 (12.8 | 73.5 (12.7 | 74.7 (11.9 | 72.2 (14.4) | 73.6 (12.8 | 74.6 (10.4 | 74.2 (13.1 | 75.1 (11.9 | 75.1 (11.9 | 75.7 (11.8 | 75.1 (11.9 | 75.8 (11.5 | 74.7 (13.2 | 74.4 (12.9 | 76.9 (14.2) |
| Carriers median age (range) (at censorship) | 75.8 (31.0-104. | 75.7 (31.0- | 75.7 (21.0- | 76.1 (37.1-1 | 74.4 (34.0- | 73.5 (34.0- | 74.5 (36.5- | 71.2 (38.1-9 | 74.2 (34.0- | 73.3 (54.8- | 76.0 (37.1- | 75.9 (31.0- | 75.7 (31.0- | 76.7 (38.8- | 75.9 (31.0- | 76.1 (31.0- | 75.2 (40.6- | 75.0 (40.6- | 79.1 (43.6-94.6 |
| **Age <40 yrs (%) (at censorship)** | 0.0087 | 0.0077 | 0.0079 | 0.0273 | 0.0158 | 0.0135 | 0.0049 | 0.0175 | 0.0174 | 0.0000 | 0.0211 | 0.0195 | 0.0074 | 0.0047 | 0.0071 | 0.0063 | 0.0000 | 0.0000 | 0.0000 |
| 40-<65 yrs (%) (at censorship) | 0.1653 | 0.1652 | 0.1661 | 0.1818 | 0.1924 | 0.1982 | 0.1650 | 0.2456 | 0.2000 | 0.1400 | 0.1789 | 0.1608 | 0.1631 | 0.1462 | 0.1610 | 0.1376 | 0.1860 | 0.16129 | 0.2000 |
| 65+yrs (%) (at censorship) | 0.8260 | 0.8271 | 0.8260 | 0.7909 | 0.7918 | 0.7883 | 0.8301 | 0.7368 | 0.7826 | 0.8600 | 0.8000 | 0.8316 | 0.8295 | 0.8491 | 0.8319 | 0.8561 | 0.8140 | 0.83871 | 0.8000 |
| Carrier Ancestry | | | | | | | | | | | | | | | | | | | |
| **EUR (%)** | 0.7741 | 0.7801 | 0.7686 | 0.6636 | 0.6814 | 0.6937 | 0.7039 | 0.6491 | 0.6783 | 0.7800 | 0.6526 | 0.7926 | 0.7767 | 0.8962 | 0.7922 | 0.8001 | 0.7209 | 0.7419 | 0.7500 |
| **AFR (%)** | 0.1643 | 0.1601 | 0.1673 | 0.2545 | 0.2492 | 0.2477 | 0.2524 | 0.2982 | 0.2348 | 0.2200 | 0.2526 | 0.1483 | 0.1624 | 0.0566 | 0.1486 | 0.1433 | 0.1860 | 0.1290 | 0.2000 |
| AMR (%) | 0.0465 | 0.0460 | 0.0488 | 0.0545 | 0.0536 | 0.0495 | 0.0340 | 0.0526 | 0.0696 | 0.0000 | 0.0632 | 0.0437 | 0.0453 | 0.0330 | 0.0438 | 0.0434 | 0.0930 | 0.1290 | 0.0500 |
| EAS | 0.0044 | 0.0041 | 0.0049 | 0.0091 | 0.0063 | 0.0045 | 0.0049 | 0.0000 | 0.0087 | 0.0000 | 0.0105 | 0.0041 | 0.0047 | 0.0000 | 0.0041 | 0.0031 | 0.0000 | 0.0000 | 0.0000 |
| SAS | 0.0005 | 0.0005 | 0.0006 | 0.0000 | 0.0000 | 0.0000 | 0.0000 | 0.0000 | 0.0000 | 0.0000 | 0.0000 | 0.0006 | 0.0007 | 0.0000 | 0.0006 | 0.0006 | 0.0000 | 0.0000 | 0.0000 |
| Other | 0.0102 | 0.0097 | 0.0098 | 0.0182 | 0.0095 | 0.0045 | 0.0049 | 0.0000 | 0.0087 | 0.0000 | 0.0211 | 0.0106 | 0.0101 | 0.0142 | 0.0107 | 0.0094 | 0.0000 | 0.0000 | 0.0000 |

***Supplementary Table ST3:*** Aggregate/summary data comparisons to UKB/GEL Stevelink data

| | All Stevelink - all variants queried | **All Stevelink - overlapping variants only** | All ClinGen overlap - all variants queried | **All ClinGen overlap - overlapping variants only** | Unclassified Genes - all variants queried | **Unclassified Genes - overlapping variants only** |
|---|---|---|---|---|---|---|
| Genes attempted | 91 | **85** | 40 | **39** | 51 | **46** |
| Variants attempted by MVP | 7112 | **2982** | 5296 | **2529** | 1815 | **453** |
| Variants attempted by UKB/GEL | 4288 | **2982** | 3612 | **2529** | 676 | **453** |
| Genes detected in MVP | 45 | **23** | 25 | **13** | 20 | **10** |
| Genes detected in UKB | 33 | **30** | 21 | **21** | 11 | **9** |
| Genes detected in GEL | 15 | **12** | 9 | **8** | 6 | **4** |
| Variants detected in MVP | 116 | **45** | 78 | **32** | 38 | **13** |
| Variants detected in UKB | 111 | **93** | 81 | **70** | 29 | **23** |
| Variants detected in GEL | 37 | **28** | 25 | **20** | 12 | **8** |
| % variants detected/attempted MVP | 0.0231 | **0.0151** | 0.0147 | **0.0127** | 0.0209 | **0.0287** |
| % variants detected/attempted UKB | 0.0679 | **0.0312** | 0.0224 | **0.0277** | 0.0429 | **0.0508** |
| % variants detected/attempted GEL | 0.0105 | **0.0094** | 0.0069 | **0.0079** | 0.0178 | **0.0177** |
| Carriers (all) in MVP | 164 | **64** | 107 | **46** | 57 | **18** |
| Carriers (all) in UKB | 291 | **230** | 239 | **194** | 52 | **36** |
| Carriers (all) in GEL | 45 | **34** | 31 | **26** | 14 | **8** |
| Affected (epilepsy) in MVP | 17 | **7** | 8 | **2** | 9 | **5** |
| Affected (epilepsy) in UKB | 9 | **9** | 9 | **9** | 0 | **0** |
| Affected (epilepsy) in GEL | 14 | **10** | 12 | **9** | 2 | **1** |
| Proportion w epi MVP (affected/carriers: aka prevalence of epilepsy among carriers) | 0.1037 | **0.1094** | 0.0748 | **0.0435** | 0.1579 | **0.2778** |
| Proportion w epi UKB (affected/carriers: aka prevalence of epilepsy among carriers) | 0.0309 | **0.0391** | 0.0377 | **0.0464** | 0.0000 | **0.0000** |
| Proportion w epi GEL (affected/carriers: aka prevalence of epilepsy among carriers) | 0.3111 | **0.2941** | 0.3871 | **0.3462** | 0.1429 | **0.1250** |
| Variant frequency in all MVP | 0.0016 | **0.0006** | 0.0010 | **0.0004** | 0.0006 | **0.0002** |
| Variant frequency in all UKB | 0.0007 | **0.0006** | 0.0006 | **0.0005** | 0.0001 | **0.0001** |
| Variant frequency in all GEL | 0.0010 | **0.0008** | 0.0007 | **0.0006** | 0.0003 | **0.0002** |
| Variant frequency in epilepsy in MVP | 0.0026 | **0.0011** | 0.0012 | **0.0003** | 0.0014 | **0.0008** |
| Variant frequency in epilepsy in UKB | 0.0113 | **0.0113** | 0.0113 | **0.0113** | 0.0000 | **0.0000** |
| Variant frequency in epilepsy in GEL | 0.0076 | **0.0054** | 0.0065 | **0.0049** | 0.0011 | **0.0005** |
| Variant frequency in non-epilepsy in MVP | 0.0015 | **0.0006** | 0.0010 | **0.0005** | 0.0005 | **0.0001** |
| Variant frequency in non-epilepsy in UKB | 0.0007 | **0.0006** | 0.0006 | **0.0005** | 0.0001 | **0.0001** |
| Variant frequency in non-epilepsy in GEL | 0.0008 | **0.0006** | 0.0005 | **0.0004** | 0.0003 | **0.0002** |

***Supplementary Table ST4:*** MVP gene-level analyses of 170 genes

| Gene | MOI | ClinGen Classification | Unique PGVs Queried | Unique PGVs Detected | % PGVs Queried / Detected | Total Carriers of PGVs | Carriers of PGVs with Epilepsy | Epilepsy Prevalence in Carriers |
|---|---|---|---|---|---|---|---|---|
| ABAT | AR | Moderate | 30 | 4 | 13.33% | 5 | 1 | 20.00% |
| ADRA2B | AD | Refuted | 1 | 0 | 0.00% | 0 | 0 | #DIV/0! |
| ALG13 | XL | Definitive | 9 | 1 | 11.11% | 1 | 0 | 0.00% |
| AP2M1 | AD | Definitive | 1 | 0 | 0.00% | 0 | 0 | #DIV/0! |
| ARX | XL | Definitive | 50 | 2 | 4.00% | 2 | 0 | 0.00% |
| ATN1 | AD | . | 7 | 0 | 0.00% | 0 | 0 | #DIV/0! |
| ATP1A2 | AD | Definitive | 97 | 7 | 7.22% | 10 | 2 | 20.00% |
| ATP1A3 | AD | . | 145 | 2 | 1.38% | 2 | 2 | 100.00% |
| ATP6V0A1 | AD | . | 10 | 1 | 10.00% | 1 | 0 | 0.00% |
| ATP6V0C | AD | . | 15 | 0 | 0.00% | 0 | 0 | #DIV/0! |
| ATP6V1A | AD | . | 23 | 0 | 0.00% | 0 | 0 | #DIV/0! |
| ATP6V1B2 | AD | . | 17 | 0 | 0.00% | 0 | 0 | #DIV/0! |
| BRAT1 | AR | Definitive | 62 | 13 | 20.97% | 30 | 0 | 0.00% |
| CACNA1A | AD | . | 326 | 12 | 3.68% | 12 | 2 | 16.67% |
| CACNA1E | AD | . | 29 | 1 | 3.45% | 1 | 0 | 0.00% |
| CACNA2D2 | AR | Moderate | 38 | 1 | 2.63% | 1 | 0 | 0.00% |
| CACNB4 | AD | Refuted | 2 | 0 | 0.00% | 0 | 0 | #DIV/0! |
| CAPRIN1 | AD | . | 15 | 0 | 0.00% | 0 | 0 | #DIV/0! |
| CASR | AD | Disputed | 186 | 11 | 5.91% | 19 | 0 | 0.00% |
| CDK19 | AD | . | 8 | 0 | 0.00% | 0 | 0 | #DIV/0! |
| CELF2 | AD | . | 8 | 0 | 0.00% | 0 | 0 | #DIV/0! |
| CERS1 | AR | Moderate | 13 | 9 | 69.23% | 204 | 17 | 8.33% |
| CHD2 | AD | Definitive | 202 | 3 | 1.49% | 5 | 1 | 20.00% |
| CHRNA2 | AD | Limited/Disputed | 3 | 0 | 0.00% | 0 | 0 | #DIV/0! |
| CHRNA4 | AD | Definitive | 5 | 1 | 20.00% | 1 | 0 | 0.00% |
| CHRNA7 | UD | Refuted | NA | 0 | #VALUE! | 0 | 0 | #DIV/0! |
| CHRNB2 | AD | Definitive | 7 | 0 | 0.00% | 0 | 0 | #DIV/0! |
| CLCN2 | AD | Refuted | 28 | 9 | 32.14% | 22 | 1 | 4.55% |
| CLN3 | AR | Definitive | 116 | 17 | 14.66% | 94 | 3 | 3.19% |
| CLN5 | AR | Definitive | 60 | 12 | 20.00% | 49 | 2 | 4.08% |
| CLN6 | AR | Definitive | 65 | 9 | 13.85% | 20 | 2 | 10.00% |
| CLN8 | AR | Definitive | 55 | 12 | 21.82% | 31 | 3 | 9.68% |
| CNTN2 | AR | Definitive | 24 | 6 | 25.00% | 29 | 3 | 10.34% |
| CNTNAP2 | AR | Definitive | 54 | 9 | 16.67% | 19 | 1 | 5.26% |
| CPA6 | AD/AR | Refuted | 1 | 1 | 100.00% | 7 | 0 | 0.00% |
| CRH | AD | Refuted | NA | 0 | #VALUE! | 0 | 0 | #DIV/0! |
| CSNK1E | AD | . | 1 | 1 | 100.00% | 1 | 0 | 0.00% |
| CSNK2B | AD | . | 73 | 0 | 0.00% | 0 | 0 | #DIV/0! |
| CSTB | AR | Definitive/moderat( | 13 | 4 | 30.77% | 79 | 2 | 2.53% |
| CTSD | AR | Definitive | 24 | 2 | 8.33% | 3 | 0 | 0.00% |
| CTSF | AR | Definitive | 18 | 7 | 38.89% | 14 | 0 | 0.00% |
| CUX2 | AD | Moderate | 3 | 0 | 0.00% | 0 | 0 | #DIV/0! |
| CYFIP2 | AD | Definitive | 20 | 1 | 5.00% | 1 | 0 | 0.00% |
| DEPDC5 | AD | Definitive | 194 | 7 | 3.61% | 8 | 0 | 0.00% |
| DNAJC5 | AD | Moderate | 2 | 0 | 0.00% | 0 | 0 | #DIV/0! |
| DNM1 | AD/AR | Definitive/moderat | 55 | 1 | 1.82% | 4 | 0 | 0.00% |
| DOCK7 | AR | Definitive | 46 | 13 | 28.26% | 26 | 0 | 0.00% |
| EEF1A2 | AD | Definitive | 40 | 2 | 5.00% | 2 | 0 | 0.00% |
| EFHC1 | AD | Refuted | NA | 0 | #VALUE! | 0 | 0 | #DIV/0! |
| EMX2 | AD | Limited | 5 | 0 | 0.00% | 0 | 0 | #DIV/0! |
| EPM2A | AR | Definitive | 32 | 13 | 40.63% | 67 | 4 | 5.97% |
| FBXO28 | AD | . | 5 | 0 | 0.00% | 0 | 0 | #DIV/0! |
| FGF12 | AD | . | 4 | 0 | 0.00% | 0 | 0 | #DIV/0! |
| FRRS1L | AR | Definitive | 16 | 6 | 37.50% | 9 | 2 | 22.22% |
| FZR1 | AD | . | 5 | 0 | 0.00% | 0 | 0 | #DIV/0! |
| GABBR2 | AD | Moderate | 14 | 1 | 7.14% | 2 | 0 | 0.00% |
| GABRA1 | AD | Definitive | 62 | 0 | 0.00% | 0 | 0 | #DIV/0! |
| GABRA2 | AD | . | 20 | 1 | 5.00% | 1 | 0 | 0.00% |
| GABRA5 | AD | . | 6 | 0 | 0.00% | 0 | 0 | #DIV/0! |
| GABRB1 | AD | Limited | 5 | 0 | 0.00% | 0 | 0 | #DIV/0! |
| GABRB2 | AD | . | 52 | 0 | 0.00% | 0 | 0 | #DIV/0! |
| GABRB3 | AD | Definitive | 84 | 1 | 1.19% | 1 | 0 | 0.00% |
| GABRD | AD | Moderate/Limited | 4 | 0 | 0.00% | 0 | 0 | #DIV/0! |
| GABRG2 | AD | Definitive | 86 | 2 | 2.33% | 2 | 0 | 0.00% |
| GLUL | AD | Moderate | 12 | 0 | 0.00% | 0 | 0 | #DIV/0! |
| GNAO1 | AD | Definitive | 82 | 0 | 0.00% | 0 | 0 | #DIV/0! |
| GNB1 | AD | . | 46 | 3 | 6.52% | 3 | 0 | 0.00% |
| GOSR2 | AR | Definitive | 15 | 5 | 33.33% | 63 | 1 | 1.59% |
| GRIN1 | AD/AR | Definitive | 101 | 3 | 2.97% | 4 | 1 | 25.00% |
| GRIN2A | AD | Definitive | 189 | 4 | 2.12% | 6 | 0 | 0.00% |
| GRIN2B | AD | Definitive | 203 | 0 | 0.00% | 0 | 0 | #DIV/0! |
| GRIN2D | AD | Definitive | 15 | 0 | 0.00% | 0 | 0 | #DIV/0! |
| GRN | AR | Definitive | 62 | 4 | 6.45% | 4 | 1 | 25.00% |
| HCN1 | AD | Definitive | 41 | 0 | 0.00% | 0 | 0 | #DIV/0! |
| HNRNPU | AD | . | 53 | 0 | 0.00% | 0 | 0 | #DIV/0! |
| IRF2BPL | AD | Definitive | 38 | 0 | 0.00% | 0 | 0 | #DIV/0! |
| ITPA | AR | Definitive | 20 | 4 | 20.00% | 14 | 1 | 7.14% |
| KCNA1 | AD | Definitive | 42 | 2 | 4.76% | 2 | 1 | 50.00% |
| KCNA2 | AD | Definitive | 52 | 1 | 1.92% | 1 | 0 | 0.00% |
| KCNA3 | AD | . | 15 | 1 | 6.67% | 1 | 1 | 100.00% |
| KCNB1 | AD | Definitive | 96 | 0 | 0.00% | 0 | 0 | #DIV/0! |
| KCNC1 | AD | Definitive | 15 | 0 | 0.00% | 0 | 0 | #DIV/0! |
| KCNC2 | AD | . | 17 | 1 | 5.88% | 1 | 0 | 0.00% |
| KCNH5 | AD | Definitive | 10 | 2 | 20.00% | 2 | 0 | 0.00% |
| KCNK4 | AD | . | 7 | 0 | 0.00% | 0 | 0 | #DIV/0! |
| KCNMA1 | AD/AR | Definitive/moderat | 28 | 4 | 14.29% | 4 | 0 | 0.00% |
| KCNQ2 | AD | Definitive | 490 | 4 | 0.82% | 4 | 1 | 25.00% |
| KCNQ3 | AD/AR | Definitive/moderat( | 42 | 1 | 2.38% | 1 | 0 | 0.00% |
| KCNT1 | AD | Definitive | 73 | 3 | 4.11% | 3 | 0 | 0.00% |
| KCNT2 | AD | . | 12 | 2 | 16.67% | 2 | 0 | 0.00% |
| KCTD7 | AR | Definitive | 22 | 6 | 27.27% | 10 | 0 | 0.00% |
| LGI1 | AD | Definitive | 38 | 2 | 5.26% | 2 | 2 | 100.00% |
| MAGI2 | AD | Refuted | 2 | 0 | 0.00% | 0 | 0 | #DIV/0! |
| MAST3 | AD | . | 8 | 0 | 0.00% | 0 | 0 | #DIV/0! |
| MEF2C | AD | . | 76 | 3 | 3.95% | 3 | 0 | 0.00% |
| MFSD8 | AR | Definitive | 92 | 19 | 20.65% | 37 | 4 | 10.81% |
| NACC1 | AD | . | 6 | 0 | 0.00% | 0 | 0 | #DIV/0! |
| NARS1 | AD | . | 13 | 10 | 76.92% | 10 | 1 | 10.00% |
| NBEA | AD | . | 57 | 0 | 0.00% | 0 | 0 | #DIV/0! |
| NCDN | AD | . | 5 | 2 | 40.00% | 2 | 1 | 50.00% |
| NECAP1 | AR | Moderate | 2 | 2 | 100.00% | 2 | 0 | 0.00% |
| NEUROD2 | AD | . | 8 | 1 | 12.50% | 1 | 0 | 0.00% |
| NHLRC1 | AR | Definitive | 26 | 4 | 15.38% | 33 | 2 | 6.06% |
| NPRL2 | AD | Definitive | 12 | 1 | 8.33% | 1 | 0 | 0.00% |
| NPRL3 | AD | Definitive | 64 | 1 | 1.56% | 1 | 0 | 0.00% |
| NR4A2 | AD | . | 25 | 1 | 4.00% | 1 | 0 | 0.00% |
| NSF | AD | . | 4 | 0 | 0.00% | 0 | 0 | #DIV/0! |
| OTUD7A | AR | Limited | 1 | 0 | 0.00% | 0 | 0 | #DIV/0! |
| PCDH19 | XL | Definitive | 219 | 9 | 4.11% | 13 | 0 | 0.00% |
| PHACTR1 | AD | . | 6 | 0 | 0.00% | 0 | 0 | #DIV/0! |
| PIGA | XL | Definitive | 29 | 0 | 0.00% | 0 | 0 | #DIV/0! |
| PIGO | AR | Definitive | 36 | 12 | 33.33% | 38 | 5 | 13.16% |
| PLCB1 | AR | Definitive | 18 | 3 | 16.67% | 3 | 1 | 33.33% |
| PNKP | AR | Definitive | 52 | 22 | 42.31% | 35 | 3 | 8.57% |
| PNPO | AR | Definitive | 31 | 17 | 54.84% | 68 | 4 | 5.88% |
| PPP3CA | AD | . | 28 | 0 | 0.00% | 0 | 0 | #DIV/0! |
| PPT1 | AR | Definitive | 106 | 25 | 23.58% | 177 | 8 | 4.52% |
| PRICKLE1 | AD/AR | Disputed | 2 | 2 | 100.00% | 5 | 0 | 0.00% |
| PRICKLE2 | AD | Limited | 3 | 1 | 33.33% | 1 | 0 | 0.00% |
| PRIMA1 | AR | Limited | 2 | 1 | 50.00% | 1 | 0 | 0.00% |
| PRRT2 | AD | Definitive | 53 | 1 | 1.89% | 1 | 0 | 0.00% |
| PURA | AD | Definitive | 94 | 1 | 1.06% | 19 | 0 | 0.00% |
| RANBP2 | AD | Moderate | 67 | 18 | 26.87% | 48 | 3 | 6.25% |
| RBFOX3 | AD | Disputed | 1 | 0 | 0.00% | 0 | 0 | #DIV/0! |
| RHOBTB2 | AD/AR | Definitive | 14 | 0 | 0.00% | 0 | 0 | #DIV/0! |
| RNF13 | AD | . | 5 | 0 | 0.00% | 0 | 0 | #DIV/0! |
| ROGDI | AR | Definitive | 30 | 9 | 30.00% | 23 | 1 | 4.35% |
| RORA | AD | . | 27 | 0 | 0.00% | 0 | 0 | #DIV/0! |
| RORB | AD | Definitive | 38 | 1 | 2.63% | 1 | 0 | 0.00% |
| RYR3 | AD | Limited | 1 | 0 | 0.00% | 0 | 0 | #DIV/0! |
| SCARB2 | AR | Definitive | 19 | 5 | 26.32% | 10 | 0 | 0.00% |
| SCN1A | AD | Definitive/strong | 1408 | 17 | 1.21% | 27 | 1 | 3.70% |
| SCN1B | AD/AR | Definitive | 20 | 4 | 20.00% | 18 | 0 | 0.00% |
| SCN2A | AD | Definitive | 517 | 5 | 0.97% | 8 | 0 | 0.00% |
| SCN3A | AD | Definitive | 28 | 2 | 7.14% | 2 | 0 | 0.00% |
| SCN8A | AD | Definitive | 288 | 3 | 1.04% | 3 | 1 | 33.33% |
| SCN9A | AD | Refuted | 108 | 26 | 24.07% | 53 | 2 | 3.77% |
| SEMA6B | AD | Definitive | 11 | 0 | 0.00% | 0 | 0 | #DIV/0! |
| SERPINI1 | AD | Definitive | 6 | 0 | 0.00% | 0 | 0 | #DIV/0! |
| SETD1A | AD | . | 31 | 4 | 12.90% | 4 | 0 | 0.00% |
| SETD1B | AD | . | 43 | 0 | 0.00% | 0 | 0 | #DIV/0! |
| SIK1 | AD | Limited | NA | - | #VALUE! | - | - | #VALUE! |
| SLC12A5 | AR | Limited | 26 | 2 | 7.69% | 2 | 0 | 0.00% |
| SLC1A2 | AD | . | 9 | 0 | 0.00% | 0 | 0 | #DIV/0! |
| SLC25A22 | AR | Definitive | 14 | 3 | 21.43% | 7 | 0 | 0.00% |
| SLC2A1 | AD | . | 197 | 4 | 2.03% | 4 | 1 | 25.00% |
| SLC32A1 | AD | . | 6 | 0 | 0.00% | 0 | 0 | #DIV/0! |
| SLC35A2 | XL | Definitive | 31 | 0 | 0.00% | 0 | 0 | #DIV/0! |
| SLC4A10 | AR | Strong | 10 | 3 | 30.00% | 4 | 0 | 0.00% |
| SLC6A1 | AD | . | 138 | 5 | 3.62% | 5 | 0 | 0.00% |
| SNAP25 | AD | . | 24 | 0 | 0.00% | 0 | 0 | #DIV/0! |
| SPTAN1 | AD | Definitive | 71 | 1 | 1.41% | 1 | 0 | 0.00% |
| SRPX2 | XL | Refuted | 2 | 0 | 0.00% | 0 | 0 | #DIV/0! |
| ST3GAL5 | AR | Definitive | 26 | 10 | 38.46% | 21 | 4 | 19.05% |
| STX1B | AD | Definitive | 39 | 1 | 2.56% | 1 | 0 | 0.00% |
| STXBP1 | AD | Definitive | 259 | 0 | 0.00% | 0 | 0 | #DIV/0! |
| SYNGAP1 | AD | Definitive | 194 | 0 | 0.00% | 0 | 0 | #DIV/0! |
| SYNJ1 | AR | Definitive | 30 | 8 | 26.67% | 27 | 3 | 11.11% |
| SZT2 | AR | Definitive | 107 | 28 | 26.17% | 66 | 8 | 12.12% |
| TANC2 | AD | . | 25 | 0 | 0.00% | 0 | 0 | #DIV/0! |
| TPP1 | AR | Definitive | 125 | 35 | 28.00% | 336 | 33 | 9.82% |
| TRA2B | AD | . | 4 | 0 | 0.00% | 0 | 0 | #DIV/0! |
| UBA5 | AR | Definitive | 24 | 2 | 8.33% | 3 | 1 | 33.33% |
| UBE3A | AD | . | 116 | 1 | 0.86% | 1 | 1 | 100.00% |
| USP25 | AD | Disputed | NA | 0 | #VALUE! | 0 | 0 | #DIV/0! |
| WASF1 | AD | . | 7 | 1 | 14.29% | 1 | 0 | 0.00% |
| WDR37 | AD | . | 8 | 0 | 0.00% | 0 | 0 | #DIV/0! |
| WDR45 | XL | Definitive | 100 | 4 | 4.00% | 4 | 0 | 0.00% |
| WWOX | AR | Definitive | 56 | 16 | 28.57% | 42 | 4 | 9.52% |
| YWHAG | AD | . | 20 | 0 | 0.00% | 0 | 0 | #DIV/0! |

***Supplementary Table ST5:*** Gene-level comparisons of MVP, UKB and GEL data for 91 overlapping genes

| Gene | MOI | ClinGen Classification | MVP: Unique PGVs Queried | MVP: Unique PGVs Detect | MVP: Total Carrier s of PGVs | MVP: Carriers of PGVs with Epileps | MVP: Epilepsy Prevalence in Carriers | MVP: % PGVs Queried / Detected | UKB/GEL : Unique PGVs Querie | UKB: Unique PGVs Detecte d | UKB: Total Carrie rs of PGV | UKB: Carriers of PGVs with Epileps | UKB: Epilepsy Prevalence in Carriers | UKB: % PGVs Queried / Detected | GEL: Unique PGVs Detecte | GEL: Total Carriers of PGV | GEL: Carriers of PGVs with Epileps | GEL: Epilepsy Prevalence in Carriers | GEL: % PGVs Queried / Detected | Total Carriers Across Cohorts | Carriers with Epilepsy Across Cohorts | Epilepsy Prevalence Across Cohorts |
|---|---|---|---|---|---|---|---|---|---|---|---|---|---|---|---|---|---|---|---|---|---|---|
| ATN1 | AD | . | 7 | 0 | 0 | 0 | #DIV/0! | 0.00% | 6 | 0 | 0 | 0 | #DIV/0! | 0.00% | 0 | 0 | 0 | #DIV/0! | 0.00% | 0 | 0 | #DIV/0! |
| ATP1A2 | AD | Definitive | 97 | 7 | 10 | 2 | 20.00% | 7.22% | 8 | 0 | 0 | 0 | #DIV/0! | 0.00% | 0 | 0 | 0 | #DIV/0! | 0.00% | 10 | 2 | 20.00% |
| ATP1A3 | AD | . | 145 | 2 | 2 | 2 | 100.00% | 1.38% | 15 | 1 | 1 | 0 | 0.00% | 6.67% | 0 | 0 | 0 | #DIV/0! | 0.00% | 3 | 2 | 66.67% |
| ATP6V0A1 | AD | . | 10 | 1 | 1 | 0 | 0.00% | 10.00% | 8 | 0 | 0 | 0 | #DIV/0! | 0.00% | 0 | 0 | 0 | #DIV/0! | 0.00% | 1 | 0 | 0.00% |
| ATP6V0C | AD | . | 15 | 0 | 0 | 0 | #DIV/0! | 0.00% | 10 | 0 | 0 | 0 | #DIV/0! | 0.00% | 0 | 0 | 0 | #DIV/0! | 0.00% | 0 | 0 | #DIV/0! |
| ATP6V1A | AD | . | 23 | 0 | 0 | 0 | #DIV/0! | 0.00% | 11 | 0 | 0 | 0 | #DIV/0! | 0.00% | 0 | 0 | 0 | #DIV/0! | 0.00% | 0 | 0 | #DIV/0! |
| ATP6V1B2 | AD | . | 17 | 0 | 0 | 0 | #DIV/0! | 0.00% | 1 | 0 | 0 | 0 | #DIV/0! | 0.00% | 0 | 0 | 0 | #DIV/0! | 0.00% | 0 | 0 | #DIV/0! |
| CACNA1A | AD | . | 326 | 12 | 12 | 2 | 16.67% | 3.68% | 212 | 16 | 28 | 0 | 0.00% | 7.55% | 7 | 7 | 0 | 0.00% | 3.30% | 47 | 2 | 4.26% |
| CACNA1E | AD | . | 29 | 1 | 1 | 0 | 0.00% | 3.45% | 12 | 0 | 0 | 0 | #DIV/0! | 0.00% | 0 | 0 | 0 | #DIV/0! | 0.00% | 1 | 0 | 0.00% |
| CAPRIN1 | AD | . | 15 | 0 | 0 | 0 | #DIV/0! | 0.00% | 2 | 0 | 0 | 0 | #DIV/0! | 0.00% | 0 | 0 | 0 | #DIV/0! | 0.00% | 0 | 0 | #DIV/0! |
| CDK19 | AD | . | 8 | 0 | 0 | 0 | #DIV/0! | 0.00% | 4 | 0 | 0 | 0 | #DIV/0! | 0.00% | 0 | 0 | 0 | #DIV/0! | 0.00% | 0 | 0 | #DIV/0! |
| CELF2 | AD | . | 8 | 0 | 0 | 0 | #DIV/0! | 0.00% | 5 | 0 | 0 | 0 | #DIV/0! | 0.00% | 0 | 0 | 0 | #DIV/0! | 0.00% | 0 | 0 | #DIV/0! |
| CHD2 | AD | Definitive | 202 | 3 | 5 | 1 | 20.00% | 1.49% | 219 | 4 | 42 | 0 | 0.00% | 1.83% | 4 | 6 | 3 | 50.00% | 1.83% | 53 | 4 | 7.55% |
| CSNK1E | AD | . | 1 | 1 | 1 | 0 | 0.00% | 100.00% | 1 | 0 | 0 | 0 | #DIV/0! | 0.00% | 0 | 0 | 0 | #DIV/0! | 0.00% | 1 | 0 | 0.00% |
| CSNK2B | AD | . | 73 | 0 | 0 | 0 | #DIV/0! | 0.00% | 1 | 0 | 0 | 0 | #DIV/0! | 0.00% | 0 | 0 | 0 | #DIV/0! | 0.00% | 0 | 0 | #DIV/0! |
| CUX2 | AD | Moderate | 3 | 0 | 0 | 0 | #DIV/0! | 0.00% | 2 | 0 | 0 | 0 | #DIV/0! | 0.00% | 0 | 0 | 0 | #DIV/0! | 0.00% | 0 | 0 | #DIV/0! |
| CYFIP2 | AD | Definitive | 20 | 1 | 1 | 0 | 0.00% | 5.00% | 16 | 0 | 0 | 0 | #DIV/0! | 0.00% | 0 | 0 | 0 | #DIV/0! | 0.00% | 1 | 0 | 0.00% |
| DEPDC5 | AD | Definitive | 194 | 7 | 8 | 0 | 0.00% | 3.61% | 252 | 14 | 20 | 3 | 15.00% | 5.56% | 5 | 5 | 3 | 60.00% | 1.98% | 33 | 6 | 18.18% |
| DNM1 | AD/AR | Definitive/moderat | 55 | 1 | 4 | 0 | 0.00% | 1.82% | 35 | 0 | 0 | 0 | #DIV/0! | 0.00% | 0 | 0 | 0 | #DIV/0! | 0.00% | 4 | 0 | 0.00% |
| EEF1A2 | AD | Definitive | 40 | 2 | 2 | 0 | 0.00% | 5.00% | 16 | 1 | 1 | 0 | 0.00% | 6.25% | 0 | 0 | 0 | #DIV/0! | 0.00% | 3 | 0 | 0.00% |
| FBXO28 | AD | . | 5 | 0 | 0 | 0 | #DIV/0! | 0.00% | 6 | 0 | 0 | 0 | #DIV/0! | 0.00% | 0 | 0 | 0 | #DIV/0! | 0.00% | 0 | 0 | #DIV/0! |
| FGF12 | AD | . | 4 | 0 | 0 | 0 | #DIV/0! | 0.00% | 2 | 0 | 0 | 0 | #DIV/0! | 0.00% | 1 | 1 | 1 | 100.00% | 50.00% | 1 | 1 | 100.00% |
| FZR1 | AD | . | 5 | 0 | 0 | 0 | #DIV/0! | 0.00% | 4 | 0 | 0 | 0 | #DIV/0! | 0.00% | 0 | 0 | 0 | #DIV/0! | 0.00% | 0 | 0 | #DIV/0! |
| GABBR2 | AD | Moderate | 14 | 1 | 2 | 0 | 0.00% | 7.14% | 9 | 1 | 1 | 0 | 0.00% | 11.11% | 0 | 0 | 0 | #DIV/0! | 0.00% | 3 | 0 | 0.00% |
| GABRA1 | AD | Definitive | 62 | 0 | 0 | 0 | #DIV/0! | 0.00% | 44 | 0 | 0 | 0 | #DIV/0! | 0.00% | 0 | 0 | 0 | #DIV/0! | 0.00% | 0 | 0 | #DIV/0! |
| GABRA2 | AD | . | 20 | 1 | 1 | 0 | 0.00% | 5.00% | 6 | 0 | 0 | 0 | #DIV/0! | 0.00% | 0 | 0 | 0 | #DIV/0! | 0.00% | 1 | 0 | 0.00% |
| GABRA5 | AD | . | 6 | 0 | 0 | 0 | #DIV/0! | 0.00% | 4 | 1 | 1 | 0 | 0.00% | 25.00% | 0 | 0 | 0 | #DIV/0! | 0.00% | 1 | 0 | 0.00% |
| GABRB1 | AD | Limited | 5 | 0 | 0 | 0 | #DIV/0! | 0.00% | 5 | 0 | 0 | 0 | #DIV/0! | 0.00% | 0 | 0 | 0 | #DIV/0! | 0.00% | 0 | 0 | #DIV/0! |
| GABRB2 | AD | . | 52 | 0 | 0 | 0 | #DIV/0! | 0.00% | 12 | 0 | 0 | 0 | #DIV/0! | 0.00% | 0 | 0 | 0 | #DIV/0! | 0.00% | 0 | 0 | #DIV/0! |
| GABRB3 | AD | Definitive | 84 | 1 | 1 | 0 | 0.00% | 1.19% | 50 | 2 | 2 | 0 | 0.00% | 4.00% | 0 | 0 | 0 | #DIV/0! | 0.00% | 3 | 0 | 0.00% |
| GABRD | AD | Moderate/Limited | 4 | 0 | 0 | 0 | #DIV/0! | 0.00% | 1 | 0 | 0 | 0 | #DIV/0! | 0.00% | 0 | 0 | 0 | #DIV/0! | 0.00% | 0 | 0 | #DIV/0! |
| GABRG2 | AD | Definitive | 86 | 2 | 2 | 0 | 0.00% | 2.33% | 69 | 0 | 0 | 0 | #DIV/0! | 0.00% | 0 | 0 | 0 | #DIV/0! | 0.00% | 2 | 0 | 0.00% |
| GNAO1 | AD | Definitive | 82 | 0 | 0 | 0 | #DIV/0! | 0.00% | 54 | 2 | 2 | 0 | 0.00% | 3.70% | 0 | 0 | 0 | #DIV/0! | 0.00% | 2 | 0 | 0.00% |
| GNB1 | AD | . | 46 | 3 | 3 | 0 | 0.00% | 6.52% | 1 | 1 | 1 | 0 | 0.00% | 100.00% | 0 | 0 | 0 | #DIV/0! | 0.00% | 4 | 0 | 0.00% |
| GRIN1 | AD/AR | Definitive | 101 | 3 | 4 | 1 | 25.00% | 2.97% | 5 | 0 | 0 | 0 | #DIV/0! | 0.00% | 0 | 0 | 0 | #DIV/0! | 0.00% | 4 | 1 | 25.00% |
| GRIN2A | AD | Definitive | 189 | 4 | 6 | 0 | 0.00% | 2.12% | 7 | 0 | 0 | 0 | #DIV/0! | 0.00% | 0 | 0 | 0 | #DIV/0! | 0.00% | 6 | 0 | 0.00% |
| GRIN2B | AD | Definitive | 203 | 0 | 0 | 0 | #DIV/0! | 0.00% | 48 | 1 | 1 | 0 | 0.00% | 2.08% | 2 | 2 | 0 | 0.00% | 4.17% | 3 | 0 | 0.00% |
| GRIN2D | AD | Definitive | 15 | 0 | 0 | 0 | #DIV/0! | 0.00% | 9 | 0 | 0 | 0 | #DIV/0! | 0.00% | 0 | 0 | 0 | #DIV/0! | 0.00% | 0 | 0 | #DIV/0! |
| HCN1 | AD | Definitive | 41 | 0 | 0 | 0 | #DIV/0! | 0.00% | 29 | 2 | 2 | 0 | 0.00% | 6.90% | 0 | 0 | 0 | #DIV/0! | 0.00% | 2 | 0 | 0.00% |
| HNRNPU | AD | . | 53 | 0 | 0 | 0 | #DIV/0! | 0.00% | 85 | 1 | 2 | 0 | 0.00% | 1.18% | 0 | 0 | 0 | #DIV/0! | 0.00% | 2 | 0 | 0.00% |
| KCNA1 | AD | Definitive | 42 | 2 | 2 | 1 | 50.00% | 4.76% | 2 | 0 | 0 | 0 | #DIV/0! | 0.00% | 0 | 0 | 0 | #DIV/0! | 0.00% | 2 | 1 | 50.00% |
| KCNA2 | AD | Definitive | 52 | 1 | 1 | 0 | 0.00% | 1.92% | 31 | 0 | 0 | 0 | #DIV/0! | 0.00% | 0 | 0 | 0 | #DIV/0! | 0.00% | 1 | 0 | 0.00% |
| KCNA3 | AD | . | 15 | 1 | 1 | 1 | 100.00% | 6.67% | 13 | 2 | 3 | 0 | 0.00% | 15.38% | 0 | 0 | 0 | #DIV/0! | 0.00% | 4 | 1 | 25.00% |
| KCNB1 | AD | Definitive | 96 | 0 | 0 | 0 | #DIV/0! | 0.00% | 58 | 0 | 0 | 0 | #DIV/0! | 0.00% | 0 | 0 | 0 | #DIV/0! | 0.00% | 0 | 0 | #DIV/0! |
| KCNC1 | AD | Definitive | 15 | 0 | 0 | 0 | #DIV/0! | 0.00% | 8 | 1 | 5 | 0 | 0.00% | 12.50% | 0 | 0 | 0 | #DIV/0! | 0.00% | 5 | 0 | 0.00% |
| KCNC2 | AD | . | 17 | 1 | 1 | 0 | 0.00% | 5.88% | 9 | 1 | 1 | 0 | 0.00% | 11.11% | 0 | 0 | 0 | #DIV/0! | 0.00% | 2 | 0 | 0.00% |
| KCNH5 | AD | Definitive | 10 | 2 | 2 | 0 | 0.00% | 20.00% | 7 | 1 | 1 | 1 | 100.00% | 14.29% | 0 | 0 | 0 | #DIV/0! | 0.00% | 3 | 1 | 33.33% |
| KCNK4 | AD | . | 7 | 0 | 0 | 0 | #DIV/0! | 0.00% | 1 | 0 | 0 | 0 | #DIV/0! | 0.00% | 0 | 0 | 0 | #DIV/0! | 0.00% | 0 | 0 | #DIV/0! |
| KCNQ2 | AD | Definitive | 490 | 4 | 4 | 1 | 25.00% | 0.82% | 411 | 10 | 20 | 2 | 10.00% | 2.43% | 3 | 3 | 2 | 66.67% | 0.73% | 27 | 5 | 18.52% |
| KCNQ3 | AD/AR | Definitive/moderat | 42 | 1 | 1 | 0 | 0.00% | 2.38% | 1 | 0 | 0 | 0 | #DIV/0! | 0.00% | 0 | 0 | 0 | #DIV/0! | 0.00% | 1 | 0 | 0.00% |
| KCNT1 | AD | Definitive | 73 | 3 | 3 | 0 | 0.00% | 4.11% | 36 | 2 | 8 | 0 | 0.00% | 5.56% | 1 | 1 | 1 | 100.00% | 2.78% | 12 | 1 | 8.33% |
| KCNT2 | AD | . | 12 | 2 | 2 | 0 | 0.00% | 16.67% | 13 | 1 | 10 | 0 | 0.00% | 7.69% | 1 | 2 | 0 | 0.00% | 7.69% | 14 | 0 | 0.00% |
| MAST3 | AD | . | 8 | 0 | 0 | 0 | #DIV/0! | 0.00% | 4 | 0 | 0 | 0 | #DIV/0! | 0.00% | 0 | 0 | 0 | #DIV/0! | 0.00% | 0 | 0 | #DIV/0! |
| MEF2C | AD | . | 76 | 3 | 3 | 0 | 0.00% | 3.95% | 2 | 0 | 0 | 0 | #DIV/0! | 0.00% | 0 | 0 | 0 | #DIV/0! | 0.00% | 3 | 0 | 0.00% |
| NACC1 | AD | . | 6 | 0 | 0 | 0 | #DIV/0! | 0.00% | 1 | 0 | 0 | 0 | #DIV/0! | 0.00% | 0 | 0 | 0 | #DIV/0! | 0.00% | 0 | 0 | #DIV/0! |
| NARS1 | AD | . | 13 | 10 | 10 | 1 | 10.00% | 76.92% | 2 | 1 | 1 | 0 | 0.00% | 50.00% | 0 | 0 | 0 | #DIV/0! | 0.00% | 11 | 1 | 9.09% |
| NBEA | AD | . | 57 | 0 | 0 | 0 | #DIV/0! | 0.00% | 23 | 0 | 0 | 0 | #DIV/0! | 0.00% | 0 | 0 | 0 | #DIV/0! | 0.00% | 0 | 0 | #DIV/0! |
| NCDN | AD | . | 5 | 2 | 2 | 1 | 50.00% | 40.00% | 4 | 0 | 0 | 0 | #DIV/0! | 0.00% | 0 | 0 | 0 | #DIV/0! | 0.00% | 2 | 1 | 50.00% |
| NEUROD2 | AD | . | 8 | 1 | 1 | 0 | 0.00% | 12.50% | 3 | 0 | 0 | 0 | #DIV/0! | 0.00% | 0 | 0 | 0 | #DIV/0! | 0.00% | 1 | 0 | 0.00% |
| NR4A2 | AD | . | 25 | 1 | 1 | 0 | 0.00% | 4.00% | 1 | 0 | 0 | 0 | #DIV/0! | 0.00% | 0 | 0 | 0 | #DIV/0! | 0.00% | 1 | 0 | 0.00% |
| NSF | AD | . | 4 | 0 | 0 | 0 | #DIV/0! | 0.00% | 2 | 0 | 0 | 0 | #DIV/0! | 0.00% | 0 | 0 | 0 | #DIV/0! | 0.00% | 0 | 0 | #DIV/0! |
| PHACTR1 | AD | . | 6 | 0 | 0 | 0 | #DIV/0! | 0.00% | 6 | 0 | 0 | 0 | #DIV/0! | 0.00% | 0 | 0 | 0 | #DIV/0! | 0.00% | 0 | 0 | #DIV/0! |
| PPP3CA | AD | . | 28 | 0 | 0 | 0 | #DIV/0! | 0.00% | 15 | 0 | 0 | 0 | #DIV/0! | 0.00% | 0 | 0 | 0 | #DIV/0! | 0.00% | 0 | 0 | #DIV/0! |
| PURA | AD | Definitive | 94 | 1 | 19 | 0 | 0.00% | 1.06% | 1 | 0 | 0 | 0 | #DIV/0! | 0.00% | 0 | 0 | 0 | #DIV/0! | 0.00% | 19 | 0 | 0.00% |
| RHOBTB2 | AD/AR | Definitive | 14 | 0 | 0 | 0 | #DIV/0! | 0.00% | 5 | 2 | 2 | 0 | 0.00% | 40.00% | 0 | 0 | 0 | #DIV/0! | 0.00% | 2 | 0 | 0.00% |
| RNF13 | AD | . | 5 | 0 | 0 | 0 | #DIV/0! | 0.00% | 3 | 0 | 0 | 0 | #DIV/0! | 0.00% | 0 | 0 | 0 | #DIV/0! | 0.00% | 0 | 0 | #DIV/0! |
| RORA | AD | . | 27 | 0 | 0 | 0 | #DIV/0! | 0.00% | 16 | 0 | 0 | 0 | #DIV/0! | 0.00% | 1 | 1 | 0 | 0.00% | 6.25% | 1 | 0 | 0.00% |
| RORB | AD | Definitive | 38 | 1 | 1 | 0 | 0.00% | 2.63% | 9 | 1 | 2 | 0 | 0.00% | 11.11% | 0 | 0 | 0 | #DIV/0! | 0.00% | 3 | 0 | 0.00% |
| SCN1A | AD | Definitive/strong | 1408 | 17 | 27 | 1 | 3.70% | 1.21% | 1337 | 18 | 60 | 1 | 1.67% | 1.35% | 5 | 5 | 2 | 40.00% | 0.37% | 92 | 4 | 4.35% |
| SCN1B | AD/AR | Definitive | 20 | 4 | 18 | 0 | 0.00% | 20.00% | 4 | 0 | 0 | 0 | #DIV/0! | 0.00% | 0 | 0 | 0 | #DIV/0! | 0.00% | 18 | 0 | 0.00% |
| SCN2A | AD | Definitive | 517 | 5 | 8 | 0 | 0.00% | 0.97% | 314 | 7 | 42 | 0 | 0.00% | 2.23% | 2 | 5 | 1 | 20.00% | 0.64% | 55 | 1 | 1.82% |
| SCN3A | AD | Definitive | 28 | 2 | 2 | 0 | 0.00% | 7.14% | 17 | 1 | 1 | 0 | 0.00% | 5.88% | 0 | 0 | 0 | #DIV/0! | 0.00% | 3 | 0 | 0.00% |
| SCN8A | AD | Definitive | 288 | 3 | 3 | 1 | 33.33% | 1.04% | 145 | 1 | 1 | 0 | 0.00% | 0.69% | 1 | 1 | 0 | 0.00% | 0.69% | 5 | 1 | 20.00% |
| SEMA6B | AD | Definitive | 11 | 0 | 0 | 0 | #DIV/0! | 0.00% | 6 | 0 | 0 | 0 | #DIV/0! | 0.00% | 0 | 0 | 0 | #DIV/0! | 0.00% | 0 | 0 | #DIV/0! |
| SETD1A | AD | . | 31 | 4 | 4 | 0 | 0.00% | 12.90% | 8 | 2 | 2 | 0 | 0.00% | 25.00% | 1 | 2 | 1 | 50.00% | 12.50% | 8 | 1 | 12.50% |
| SETD1B | AD | . | 43 | 0 | 0 | 0 | #DIV/0! | 0.00% | 1 | 0 | 0 | 0 | #DIV/0! | 0.00% | 0 | 0 | 0 | #DIV/0! | 0.00% | 0 | 0 | #DIV/0! |
| SLC1A2 | AD | . | 9 | 0 | 0 | 0 | #DIV/0! | 0.00% | 7 | 0 | 0 | 0 | #DIV/0! | 0.00% | 0 | 0 | 0 | #DIV/0! | 0.00% | 0 | 0 | #DIV/0! |
| SLC2A1 | AD | . | 197 | 4 | 4 | 1 | 25.00% | 2.03% | 11 | 0 | 0 | 0 | #DIV/0! | 0.00% | 0 | 0 | 0 | #DIV/0! | 0.00% | 4 | 1 | 25.00% |
| SLC32A1 | AD | . | 6 | 0 | 0 | 0 | #DIV/0! | 0.00% | 2 | 0 | 0 | 0 | #DIV/0! | 0.00% | 0 | 0 | 0 | #DIV/0! | 0.00% | 0 | 0 | #DIV/0! |
| SLC6A1 | AD | . | 138 | 5 | 5 | 0 | 0.00% | 3.62% | 94 | 2 | 2 | 0 | 0.00% | 2.13% | 1 | 1 | 0 | 0.00% | 1.06% | 8 | 0 | 0.00% |
| SNAP25 | AD | . | 24 | 0 | 0 | 0 | #DIV/0! | 0.00% | 17 | 0 | 0 | 0 | #DIV/0! | 0.00% | 0 | 0 | 0 | #DIV/0! | 0.00% | 0 | 0 | #DIV/0! |
| SPTAN1 | AD | Definitive | 71 | 1 | 1 | 0 | 0.00% | 1.41% | 65 | 4 | 15 | 0 | 0.00% | 6.15% | 2 | 3 | 0 | 0.00% | 3.08% | 19 | 0 | 0.00% |
| STX1B | AD | Definitive | 39 | 1 | 1 | 0 | 0.00% | 2.56% | 45 | 2 | 3 | 0 | 0.00% | 4.44% | 0 | 0 | 0 | #DIV/0! | 0.00% | 4 | 0 | 0.00% |
| STXBP1 | AD | Definitive | 259 | 0 | 0 | 0 | #DIV/0! | 0.00% | 224 | 4 | 8 | 2 | 25.00% | 1.79% | 0 | 0 | 0 | #DIV/0! | 0.00% | 8 | 2 | 25.00% |
| SYNGAP1 | AD | Definitive | 194 | 0 | 0 | 0 | #DIV/0! | 0.00% | 8 | 0 | 0 | 0 | #DIV/0! | 0.00% | 0 | 0 | 0 | #DIV/0! | 0.00% | 0 | 0 | #DIV/0! |
| TANC2 | AD | . | 25 | 0 | 0 | 0 | #DIV/0! | 0.00% | 2 | 0 | 0 | 0 | #DIV/0! | 0.00% | 0 | 0 | 0 | #DIV/0! | 0.00% | 0 | 0 | #DIV/0! |
| TRA2B | AD | . | 4 | 0 | 0 | 0 | #DIV/0! | 0.00% | 1 | 0 | 0 | 0 | #DIV/0! | 0.00% | 0 | 0 | 0 | #DIV/0! | 0.00% | 0 | 0 | #DIV/0! |
| UBE3A | AD | . | 116 | 1 | 1 | 1 | 100.00% | 0.86% | 1 | 0 | 0 | 0 | #DIV/0! | 0.00% | 0 | 0 | 0 | #DIV/0! | 0.00% | 1 | 1 | 100.00% |
| WASF1 | AD | . | 7 | 1 | 1 | 0 | 0.00% | 14.29% | 1 | 0 | 0 | 0 | #DIV/0! | 0.00% | 0 | 0 | 0 | #DIV/0! | 0.00% | 1 | 0 | 0.00% |
| WDR37 | AD | . | 8 | 0 | 0 | 0 | #DIV/0! | 0.00% | 1 | 0 | 0 | 0 | #DIV/0! | 0.00% | 0 | 0 | 0 | #DIV/0! | 0.00% | 0 | 0 | #DIV/0! |
| YWHAG | AD | . | 20 | 0 | 0 | 0 | #DIV/0! | 0.00% | 4 | 0 | 0 | 0 | #DIV/0! | 0.00% | 0 | 0 | 0 | #DIV/0! | 0.00% | 0 | 0 | #DIV/0! |
